# Modifiers of Influenza Vaccine Immunity from over Ten Years of Serological Data

**DOI:** 10.64898/2026.01.07.25342310

**Authors:** Emilio Fenoy, Ewan P. Plant, Hang Xie, Zhiping Ye, Arush Tagade, Jessica Rumbelow, Tal Einav

## Abstract

Annual influenza vaccination is the cornerstone for seasonal protection, yet antibody responses are highly variable across individuals and over time. To systematically assess the determinants of this heterogeneity, we compiled 21,373 hemagglutination inhibition and neutralization titers from 4,453 participants enrolled in 14 new vaccine studies we conducted and 50 prior studies that collectively span 2010-2023. Seasonality, age, vaccine brand, and pre-vaccination antibody titers were the largest and most consistent modifier of post-vaccination titers, outweighing the influence of sex, passaging, or vaccination history. Titers to B Yamagata remained steady throughout all years examined, suggesting unique durability and a potential reason for lineage extinction. Vaccine timing emerged as a strong and previously underappreciated determinant of immunity, with individuals vaccinated later in the season exhibiting larger post-vaccination titers. Not being vaccinated or receiving the live-attenuated FluMist vaccine significantly enhanced the response to inactivated vaccines during the following season in 53% or 67% of cohorts, respectively, whereas high-dose vaccines increased post-vaccination titers after adjusting for age and seasonality. These findings quantify how age shapes the influenza antibody responses in unprecedented resolution, identify vaccine timing and seasonal context as underrecognized drivers of immunogenicity, and provide actionable insights for optimizing influenza vaccination strategies.

## Introduction

Influenza continues to cause substantial global morbidity and mortality, and vaccination remains the most effective strategy to elicit protection. Among the immune pathways engaged by vaccination, the antibody response serves as a primary line of defense, and hemagglutination inhibition (HAI) titers are widely used as a regulatory correlate of protection.^1–3^

HAI titers exhibit striking heterogeneity across individuals and seasons, making it challenging to discern consistent patterns.^4–9^ Most prior studies enrolled 10-100 participants within a single season, providing valuable but inherently limited snapshots of immune dynamics. Multi-year cohorts spanning 3-5 years have now expanded this viewpoint, yet analyses have rarely assessed longer time spans.^10–15^ As a result, the contributions of repeated vaccination, seasonality, or host factors have not been systematically studied at scale, and the field lacks a clear framework for predicting how the population’s vaccine responsiveness changes over time. Quantifying these effects has become especially timely in the wake of the COVID-19 pandemic, which disrupted influenza circulation, vaccination patterns, and caused the extinction of the B Yamagata lineage.^16–18^ Simultaneously examining pre- and post-pandemic seasons provides a unique opportunity to quantify how these events shaped influenza antibody responses.

This work combines 14 new influenza vaccine studies with 50 existing studies to assess antibody measurements from over 4,400 individuals spanning the past decade. We used this dataset to disentangle the effects of host features (age, sex, prior exposure) and season-to-season variability. The diversity of pooled cohorts also enables a comparison of immunogenicity across current influenza vaccine brands including standard-dose inactivated influenza vaccines (IIVs such as Fluzone, Fluarix, Vaxigrip, and Afluria; 15μg antigen/viral strain), high-dose IIV (Fluzone High-Dose, 60μg antigen/viral strain), an adjuvanted vaccine (Fluad; 15μg antigen/viral strain + MF59 adjuvant),^2,19^ cell-grown vaccine (Flucelvax; 15μg antigen/viral strain), recombinant vaccine solely containing hemagglutinin (Flublok; 45μg antigen/viral strain), and a live-attenuated influenza vaccine (LAIV FluMist; 10^6^.^5–7^.^5^ fluorescent focus units of reassortants/viral strain). Whereas all other vaccines are administered intramuscularly, FluMist is a nasal spray and does not typically elicit a systemic antibody response in adults.^20–22^ In theory, these vaccines could trigger different immune pathways and lead to distinct antibody responses, but few analyses compared their immunogenicity across multiple seasons.^23,24^

Regarding vaccination history, prior work showed that more frequent vaccinations are associated with attenuated antibody responses.^9,25–30^ In those studies, the biggest decline in antibody titers was seen between groups that received 0 vs 1 total vaccinations anytime in the past 5 years, with more gradual decreases seen between groups vaccinated 1-5 times in that same period.^9,28^ However, it remains unclear how the number of years examined affects this association, nor whether more recent years have more impact than earlier vaccinations.

Lastly, we examine how vaccine brand or the timing of vaccination across the influenza season affects antibody titers. At present, organizations such as the WHO’s Strategic Advisory Group or the Centers for Disease Control and Prevention (CDC) list multiple options for influenza vaccines but offer few guidelines to choose among them.^31,32^ Aside from several age restrictions (*e.g.*, high-dose or adjuvanted vaccines recommended for ≥65 y.o., recombinant vaccines for ≥9 y.o., live-attenuated vaccines for 2-49 y.o.^33^), current recommendations state that most healthy individuals 6 months or older should receive an annual influenza vaccine^34^ but minimally comment upon the timing of vaccine administration^34–36^ nor suggest changes based on recent vaccination or infection history.^37^ By assessing the vaccine options made across numerous studies, this work aims to uncover actionable practices that elicit the best possible antibody response.

## Results

### Intra-study variance is more than 10x larger than cross-study variance, suggesting that datasets can be jointly analyzed

Given that the drivers of vaccine-induced immunity may be highly interdependent, we first characterized post-vaccination antibody titers as a function of all captured host, virus, and vaccine features using a multivariable linear mixed-effects model. Analysis of the variance components yielded an intra-class correlation of 7.9% (cross-dataset variance = 0.13; within-dataset residual variance = 1.52). This low intra-class correlation quantitatively demonstrated that the variance in titers was far greater within each cohort than between cohorts, suggesting that there were no systematic biases across studies. Hence, data from multiple studies were pooled in all subsequent analyses.

Functional antibody titers are commonly measured using hemagglutination inhibition (HAI) or neutralization (Neut). Despite the differences between these assays, cases where HAI and neutralization were both measured in the same individuals showed minimal differences across assays (**Figure S1**). Multivariate mixed-effects modeling further showed no significant difference in log_2_(post-vaccination titers) measured by neutralization vs HAI (effect size: -0.1, p = 0.69, Wald Z-tests used unless otherwise noted), as corroborated by multiple prior studies showing strong correlation between both assays.^27,38–45^ Hence, HAI and neutralization titers (henceforth referred to as “antibody titers”) were also combined in subsequent analyses. However, neutralization measurements constituted ∼8% of all data, and when these data were removed, all of the trends reported below continued to hold.

In total, 21,373 antibody titers were combined from 4,453 participants enrolled in 64 influenza vaccine studies spanning the 2010 to 2023 seasons (**Table S1**). In all studies, titers were measured pre-vaccination and 1-month post-vaccination against that season’s vaccine strains (**Table S2**).

### Multivariate modeling of post-vaccination titers revealed seasonality, age, pre-vaccination titers, and receipt of LAIV/high-dose IIV as key determinants of immunity

Seasonality was one of the strongest determinants of post-vaccination titers identified by the linear mixed-effects model (**Figure 1**). Comparing titers from each season to the earliest 2010 vaccine studies showed a significant non-linear decrease in post-vaccination titers in nearly every subsequent year after adjusting for host, vaccine, and baseline titer differences, reaching its lowest point in 2023 (−2.06, p<0.05).

**Figure 1.**
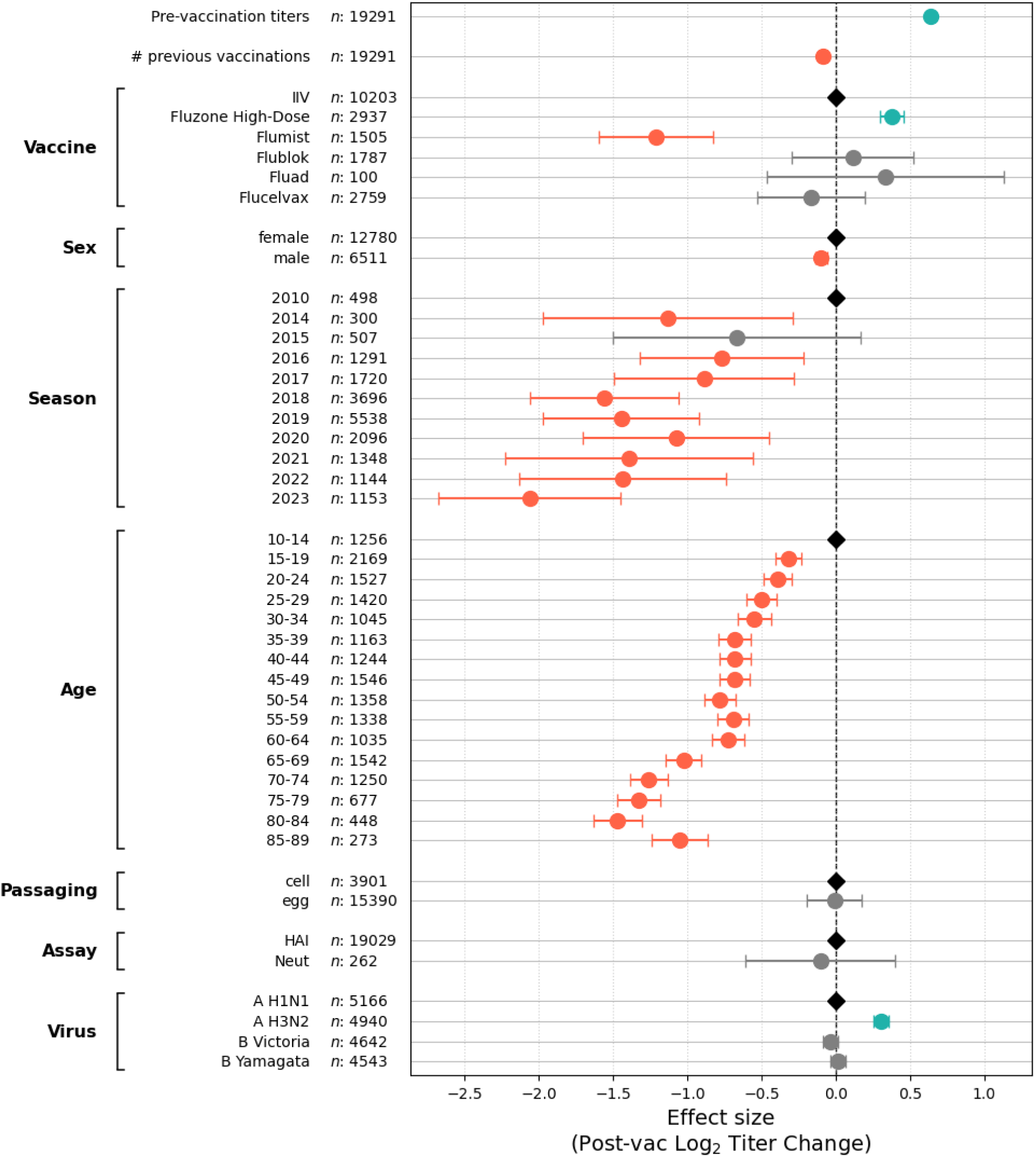
Predictors of post-vaccination titers through multivariate mixed-effects modeling. Coefficients and 95% CI of log_2_(post-vaccination titer) effect size for each feature across all datasets, with each factor having the additive linear effect shown on the *x* axis (**Methods**). *Pre-vaccination titer*, *Sex*, and *Age* denote host-specific traits. *Previous vaccination* denotes the number of consecutive prior seasons a participant was vaccinated for influenza. *Vaccine* represents the brand received, *Season* the year of vaccine administration, *Passaging* denotes if the test virus for the assay was egg- or cell-grown, *Assay* distinguishes hemagglutination inhibition (HAI) from microneutralization (Neut) titers, and *Virus* represents the subtype or lineage tested. Teal, gray, and orange circles denote features that significantly increased, were non-significant, or significantly decreased post-vaccination titers, respectively, compared to the anchoring variable (black diamond, shown when relevant).

Larger pre-vaccination titers were also positively associated with larger post-vaccination titers, with every 1-unit increase in log_2_(pre-vaccination titer) increasing log_2_(post-vaccination titer) by 0.64 units (p<0.05). Chronological age had a non-linear negative association with post-vaccination titer, with all age groups showing significantly reduced titers compared to the 10-year-old baseline (p<0.05 for all). The log_2_(post-vaccination titers) declined progressively until age 35, remained tightly clustered from ages 35-64, and then displayed the poorest and most variable values for elderly subjects 65+ years old. These trends were highly robust to both finer and coarser age binning (**Figure S2**). Interestingly, the oldest 85+ years old subjects showed *higher* titers than 80-84 year olds (relative effect size=0.42 log_2_(titer) units).

Elderly subjects receiving Fluzone High-Dose had post-vaccination titers significantly increased by +0.38 log_2_ units (p<0.05), making them on-par with subjects 35-64 years old. The adjuvanted vaccine Fluad, recombinant Flublok, and cell-grown Flucelvax led to changes of +0.34, +0.12, and -1.21 units, respectively, but none were statistically significant due to their large variability (Fluad p = 0.41, Flublok p = 0.21, Flucelvax p = 0.19). FluMist was the only vaccine with significantly lower serum titers (−1.21 units, p<0.05).

Across the other features captured in these studies, the cumulative number of prior influenza vaccinations in the dataset changed log_2_(post-vaccination titer) by -0.09 units per prior vaccination (p<0.05). Male sex was associated with a slight but statistically significant reduction of -0.1 units in log_2_ titers compared to females (p<0.05). The mixed-effects model showed no significant difference between cell-grown and egg-grown reference antigens (−0.01, p = 0.92). Titers to H3N2 were systematically higher (+0.31 units, p<0.05) than the H1N1 reference, even though B Victoria and B Yamagata were comparable to H1N1.

Building upon this multivariate analysis, we next examined the determinants of influenza immunity through univariate analyses to investigate more granular and complex trends that are difficult to isolate within a high-dimensional multivariate framework. When possible, each determinant was assessed across multiple seasons to ensure that results were robust and generalizable.

### Pre- and post-vaccination antibody titers varied substantially across seasons and showed a pronounced decline in 2023

Since the mixed-effects model identified seasonality as one of the most impactful determinants of immunogenicity, we first examined how pre-vaccination titers, post-vaccination titers, and fold-change (post-vaccination titer/pre-vaccination titer) varied across seasons. Pre-vaccination titers were remarkably stable from 2014-2018 but showed a pronounced decrease from 2021-2023 following the COVID-19 pandemic.^46,47^ Among the influenza vaccine components, H1N1 showed the most pronounced decline in pre-vaccination titers that started in 2019 (before which GMT≈40), reaching and maintaining GMT≲15 from 2021-2023 (**Figure 2A**). H3N2 and B Victoria followed this trend more gradually, dropping to comparable levels by 2022. Interestingly, B Yamagata titers did not show a sustained decline despite the lineage’s extinction during the COVID-19 lockdown (although it remained in the quadrivalent vaccine through 2023).^17,18^

**Figure 2.**
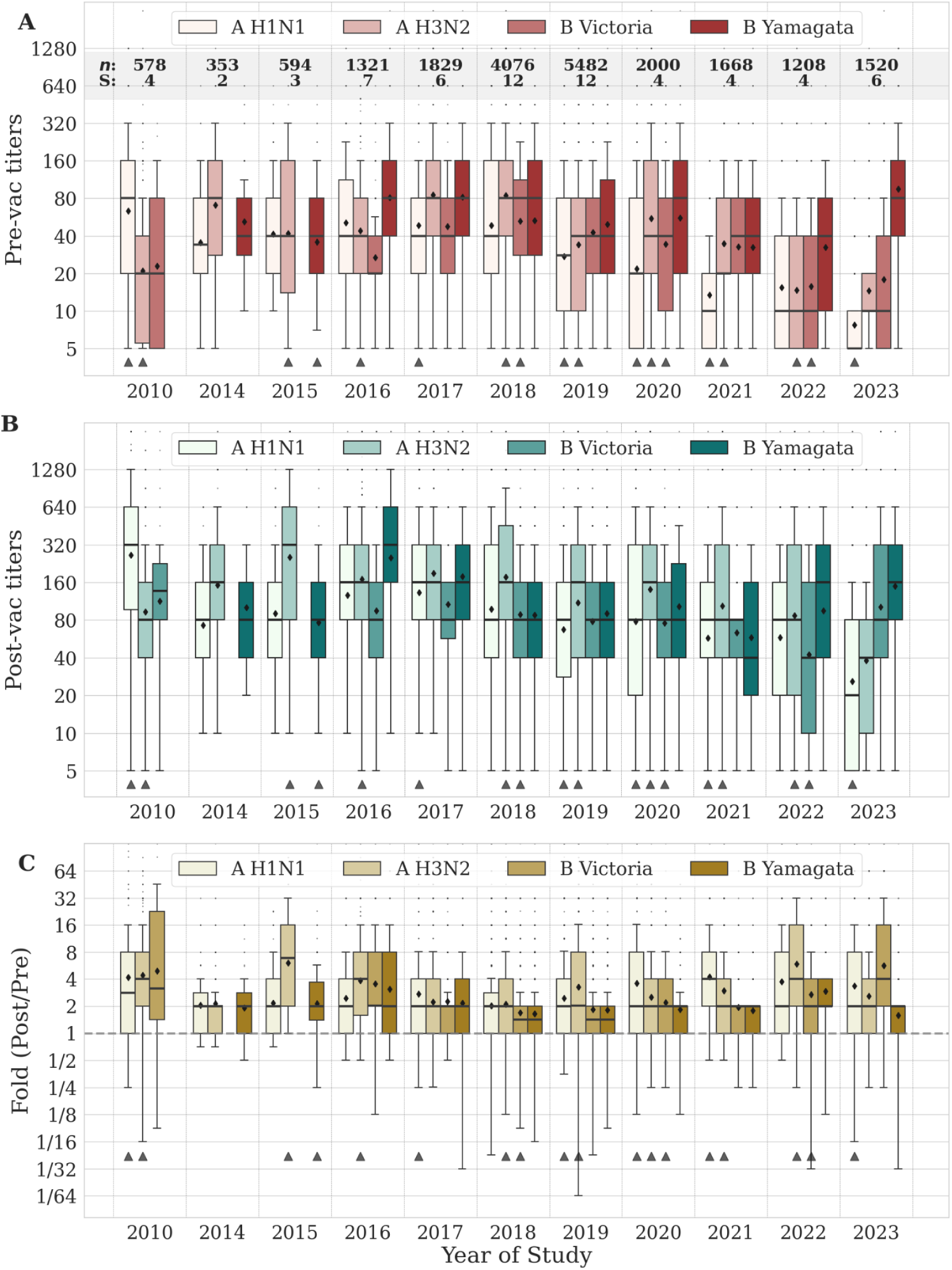
Unadjusted antibody responses to influenza vaccination across 11 seasons. (A) Pre-vaccination, (B) post-vaccination, or (C) fold-change in antibody titer across seasons by vaccine component. Panel A shows the number of studies (S) and number of measurements (*n*) analyzed. The horizontal dashed line in Panel C indicates a fold-change of 1, corresponding to no change in titer. In all univariate analyses, titers are combined across all host traits (*e.g.*, age, sex, vaccine history) unless otherwise noted. All box plots show the interquartile range, horizontal lines the medians, and black diamonds the geometric mean titers (GMTs). Whiskers extend to 1.5 times the interquartile range. Black triangles below each distribution indicate that the vaccine strain changed that season (**Table S2**).

To control for potential confounders, we fit an additional multivariate mixed-effects model on log_2_(pre-vaccination titers) (**Figure S3A**). Compared to H1N1, the other three subtypes and lineages had significantly higher pre-vaccination titers: H3N2 (coefficient = 0.33, 95% CI [0.26, 0.4], p<0.05) and B Victoria (coefficient = 0.32, 95% CI [0.25, 0.39], p<0.05) were slightly elevated while B Yamagata was noticeably larger (coefficient = 0.79, 95% CI [0.72, 0.86], p<0.05). Unlike the post-vaccination results described above, pre-vaccination titers decreased nearly monotonically from the 10-35 y.o. and then stayed relatively flat for older age groups, while male sex (coefficient = 0.07, 95% CI [0.02, 0.12], p<0.05) and a higher number of previous vaccinations (coefficient = 0.09, 95% CI [0.06, 0.11], p<0.05) were associated with marginally higher baseline titers.

Post-vaccination titers (**Figure 2B**) remained generally stable despite seasonal variation, with the exception of a distinct decrease in influenza A subtype titers in 2023. The fold-change in antibody titers also remained stable, consistently ranging between 2-4x across most strains and seasons (**Figure 2C**).

### Season-to-season variability outweighed age or consecutive vaccination effects

As suggested by the multivariate analysis, the differences in pre- and post-vaccination titers could be partly attributed to participants being older or having more repeated vaccinations. To further test these hypotheses, we first tracked antibody responses in individuals with two consecutive vaccinations across seasons, classifying them based on whether their post-vaccination antibody titers were weak (≤40) or strong (>40) against that season’s vaccine strain (*e.g.*, strong-weak means that post-vaccination titers were >40 in one season and ≤40 the next). Prior studies have found that a titer threshold of 40 roughly correlated with 50% protection,^48–52^ so that strong-strong and weak-weak represent consistently robust/frail phenotypes, whereas strong-weak or weak-strong represent a change across consecutive seasons.

The fraction of participants in each group revealed a steady decline among strong-strong responders that have been replaced by weak-weak responders (**Figure 3A**). This shift started in the 2017-2018 seasons where 18% of participants went from a strong to a weak response, while only 1% improved from weak to strong. At this inflection point, the previously dominant strong-strong group comprising ∼60% of participants began to decline, and weak-weak responses became the dominant phenotype in 2021-2022. These trends started before the COVID-19 pandemic, and they have persisted into 2022-2023.

**Figure 3.**
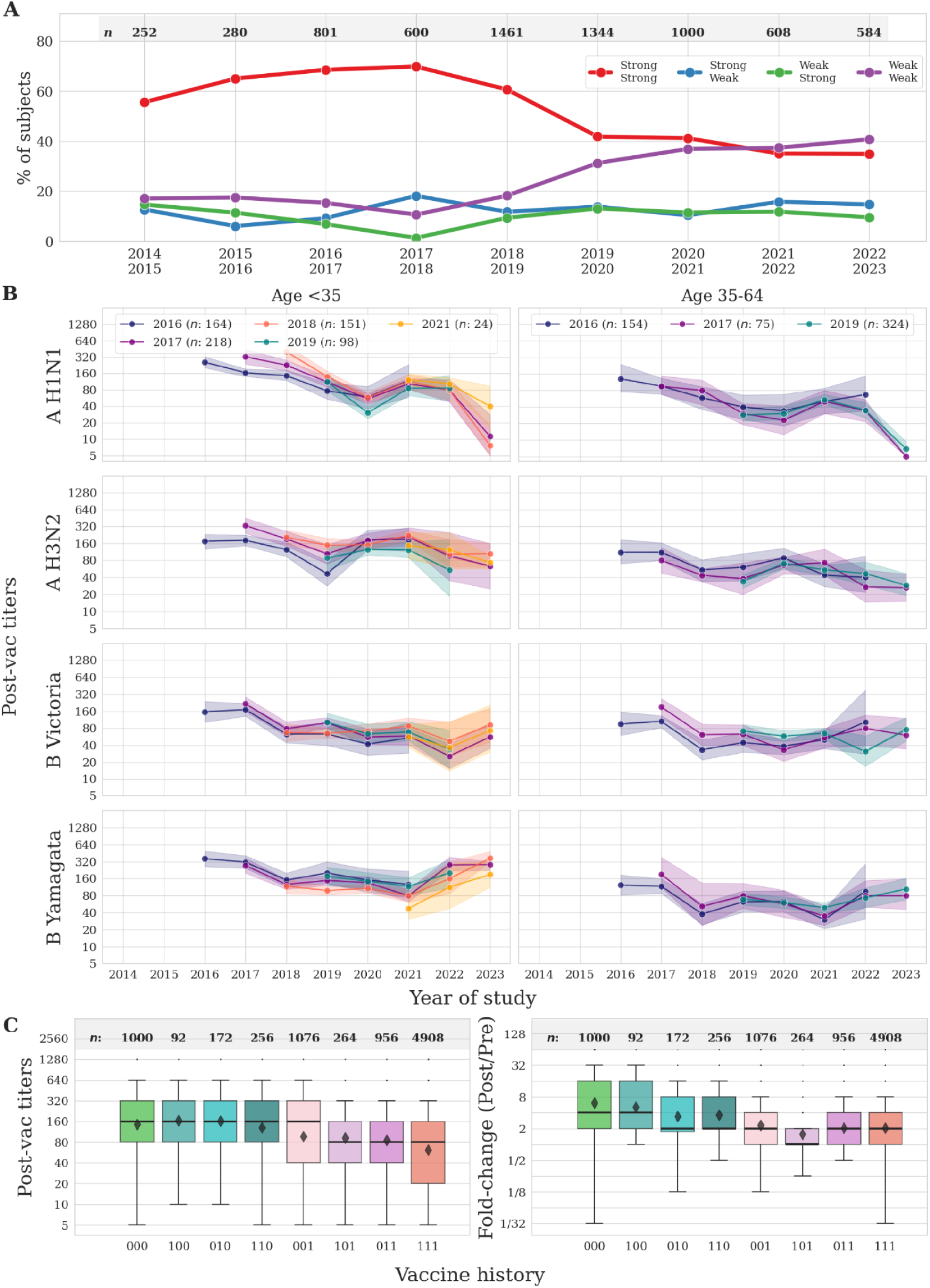
Changes in post-vaccination antibody titers for consecutively vaccinated individuals. (A) Antibody titers were categorized as weak (≤40) or strong (>40) for participants vaccinated in two consecutive seasons (*x* axis, *e.g.*, 2017-2018 implies vaccination in 2017 and 2018, irrespective of vaccines in other seasons). Percent of participants in each category (*y* axis) was computed across all vaccine strains and vaccine histories. (B) GMT of participants consecutively vaccinated in ≥3 consecutive seasons, subset by age and by the first season of their consecutive vaccines (**Methods**). Lines show GMTs and 95% confidence intervals. (C) Boxplots comparing post-vaccination antibody titers and fold-change from baseline for individuals with different vaccination histories over the preceding three years (0 = not vaccinated, 1 = vaccinated, tuples represent vaccine history [3 years ago, 2 years ago, 1 year ago]). GMTs are averaged across all seasons. The number of measurements (*n*) is shown in each panel.

Based on the multivariate analysis (**Figure 1**), we further classified participants as immunologically *young* (<35 y.o., with the highest post-vaccination titers), immunologically *adult* (35-64 y.o., intermediate titers) or immunologically *elderly* (≥65 y.o., lowest titers). Repeating the consecutive vaccination analysis in each group showed that weak-weak responses became dominant even earlier in adults and the elderly (2019-2020 and 2017-2018 respectively), while strong-strong responses have remained dominant in the young but are also trending towards a dominant weak-weak phenotype (**Figure S4A**).

To assess the impact of more frequently vaccinated individuals, we next analyzed participants that received a vaccine in three or more consecutive seasons. Antibody titers were stratified by age, vaccine component, and by the first year they participated in these consecutive vaccine studies (**Figure 3B**, **Methods**; elderly in **Figure S4B**). Titers were highly comparable across all groups. For example, H1N1 GMTs for 35-64 y.o. in 2019 ranged between 20-36 regardless of whether this was their 1st (teal), 3rd (purple), or 4th (dark blue) vaccination (**Figure 3B**).

### Individuals vaccinated in the prior season elicited 2x smaller post-vaccination titers

While the multivariate model showed that the number of consecutive prior vaccinations slightly decreases post-vaccination titers (**Figure 1**), it is unclear whether more recent years have a larger impact. Across most datasets, vaccination history was available for the past three years, which we encoded as a binary string showing either no vaccination ‘0’ or vaccination ‘1’ (*e.g.*, ’011’ means no vaccine 3 years ago followed by vaccination 2 years ago and also 1 year ago).

Individuals with no vaccinations in the preceding three years (’000’) had 2.4-fold higher post-vaccination titers and 3.0x larger fold-change than individuals vaccinated consecutively for all three years (’111’) (*p*<0.05, Welch’s t-test) (**Figure 3C**). By considering all vaccination paths, the most recent season had the strongest effect, with individuals not vaccinated in the prior season (’100’, ’010’, and ’110’) displaying post-vaccination titers statistically comparable to the vaccination-naïve (’000’) group. In contrast, participants with only one prior vaccine from the prior season (’001’), showed a significant decrease in titers compared to the naïve group (*p*<0.05, Welch’s t-test). A *de novo* mixed-effects model incorporating each prior vaccination path supported these findings. Post-vaccination titers showed large, significant decreases for subjects vaccinated in the prior year (’001’: coefficient = -0.74 95% CI [-0.84, -0.67], ’011’: coefficient = -1.04 95% CI [-1.15, -0.937], ’101’: coefficient = -1.21 95% CI [-1.37, -1.05], ’111’: coefficient = -1.14 95% CI [-1.22, -1.05]), although subjects only vaccinated two years ago elicited a smaller but still significant decrease (’010’: coefficient = -0.40 95% CI [-0.58, -0.22], ’110’: coefficient = -0.41 95% CI [-0.58, -0.25]) (**Figure S3B**). This phenomenon was highly robust and persisted after stratifying by age, season, pre-vaccination titer, and vaccine strain (**Figure S5**). Together, these results suggested that the prior year’s vaccination status was the primary driver of the effect, while the effect from vaccinations in earlier seasons exerted a smaller impact.

### Long-term antibody titers for all vaccine strains tend to remain within 25% of baseline

A different mechanism by which repeated vaccinations could impact antibody titers is by eliciting durable antibody responses that increase a subject’s pre-vaccination titers in subsequent seasons. This effect was tested in two groups: 1) participants vaccinated in two consecutive seasons and 2) participants who skipped vaccination for one season but were vaccinated in the preceding and following seasons. In the first group, pre-vaccination titers in the second season represented the day 365 titers from the prior season. In the second group, pre-vaccination titers after the skipped season represented the day 730 post-vaccination titers from the previous vaccine. In both cases, day 0 represented the pre-vaccination titer from the prior season, and later time points were always measured against the day 0 vaccine strain even if the vaccine was updated in later seasons (**Figure 4A**).

**Figure 4.**
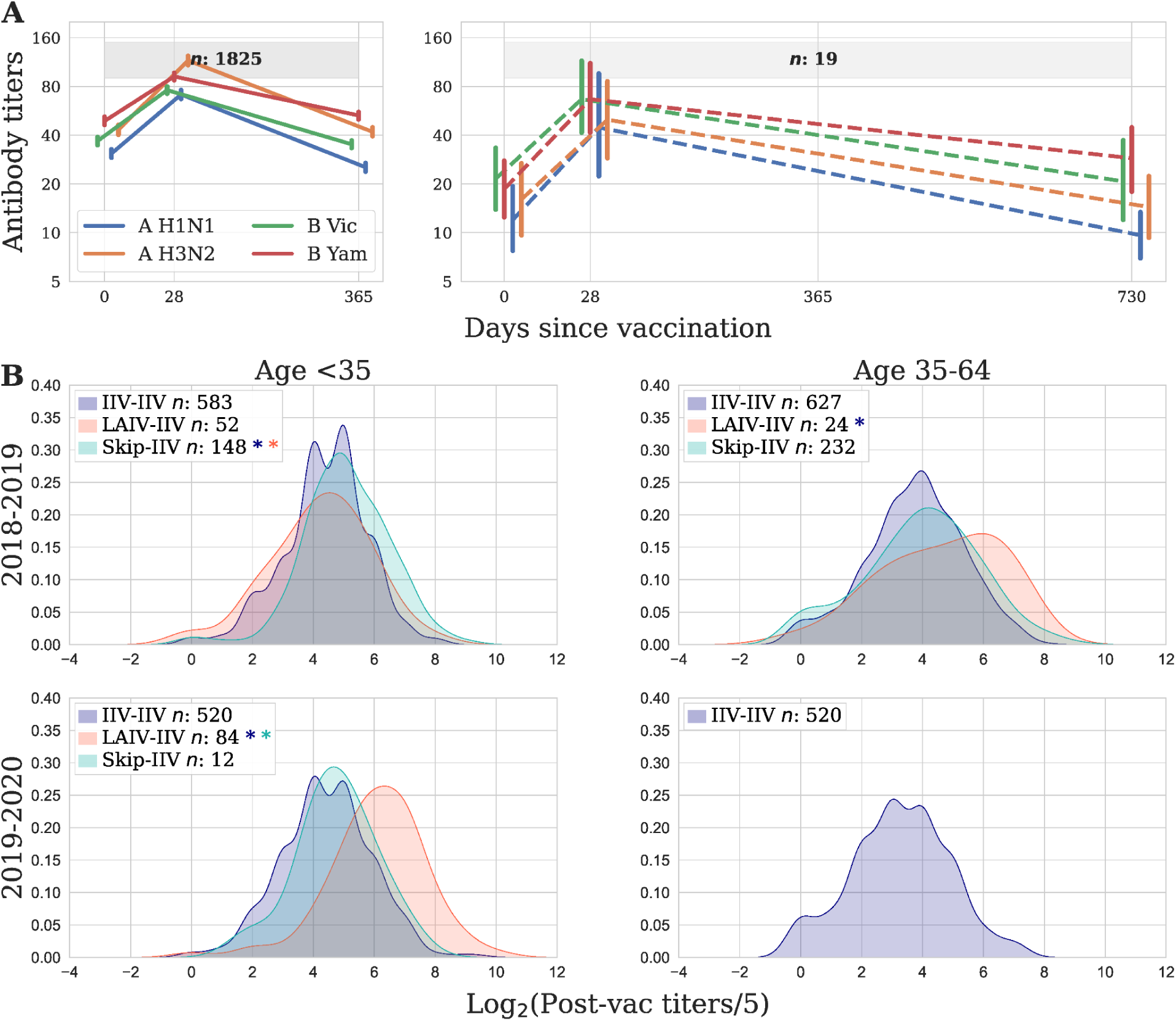
Effects of skipped vaccination on the magnitude and durability of antibody titers. (A) Long-term titers of individuals vaccinated in two consecutive years (left panel) and individuals that skipped a year in between vaccines (right panel). Antibody titers were measured at days 0, 28, 365, and 730 against the same day 0 variant, with GMTs shown across all participants and seasons. Vertical bars denote the 95% confidence intervals, and *x* coordinates are jittered for clarity. (B) Density plots of the log_2_ post-vaccination titers (titer = 5 → 0, titer = 10 → 1…) of young (<35 y.o.) and middle-aged adults (35-64 y.o.). Titers are shown for participants who received Fluzone or Flucelvax [IIV] in consecutive years (purple), no vaccination [Skip] followed by IIV (teal), or FluMist [LAIV] followed by IIV (red). A colored asterisk represents a statistically significant increase in post-vaccination titers over the group shown by that color using a Mann-Whitney U test. The number of measurements (*n*) is shown in each panel.

At day 365, antibody titers relative to day 28 markedly differed across vaccine strains, with a slower decrease for B Yamagata and B Victoria that reached 58% and 48% of their day 28 titers compared to 36% and 39% for H1N1 and H3N2. Relative to day 0, all four vaccine components remained within 15% of baseline values, with mild increases seen for both B viruses and H3N2. Thus, antibody responses showed little-to-no durability at the population-level.

At day 730, titer levels relative to day 28 moderately decreased (B Yamagata = 52%, B Victoria = 32%, H1N1 = 21%, and H3N2 = 27%). Relative to day 0, the H1N1, H3N2, and B Victoria titers remained within 25% of their baseline values, although B Yamagata showed a 68% elevation in titers at day 730 for this smaller set of participants. Hence, B Yamagata may exhibit a distinct durability compared to the other vaccine strains, although this was not seen in the far larger sampling of day 365 responses.

### Two types of “skipped” vaccination were associated with higher post-vaccination titers

Across all datasets, individuals that skipped vaccination in the prior season and then received an inactivated influenza vaccine (Skip-IIV) had significantly larger post-vaccination titers than individuals vaccinated in both seasons (IIV-IIV) in 53% = 8/15 of seasons and age groups (**Figure S6**) with no instances of the opposite trend. Prior work suggested that antibodies may mediate this effect by blocking the new vaccine’s antigen or by focusing the response towards previously-seen epitopes.^53–56^

If this is an antibody effect, then we hypothesized that another way to “skip” vaccination is to receive the FluMist vaccine, which typically does not induce a systemic antibody response.^15,57,58^ Thus, we expected individuals receiving FluMist in 2018 and IIV in 2019 (LAIV-IIV) would behave identically to the Skip-IIV group. Interestingly, while participants younger than 35 y.o. showed a statistically similar response from LAIV-IIV and IIV-IIV, middle-aged adults (35-64 y.o.) had significantly higher titers from LAIV-IIV than from IIV-IIV, suggesting that FluMist enhanced the subsequent IIV response (**Figure 4B**).^22,59^ In the subsequent 2019-2020 seasons, <35 y.o. subjects showed significantly larger titers from LAIV-IIV than IIV-IIV or Skip-IIV (**Figure 4B**, LAIV-IIV not given to 35-64 y.o. in 2019-2020). While preliminary, these results showed enhanced responses in 67% = 2/3 of the seasons and age groups examined. These effects were consistent with a *de novo* mixed-effects model specifically designed to compare IIV-IIV, Skip-IIV, and LAIV-IIV, where Skip-IIV was associated with a significant 1.06 log_2_-unit increase in post-vaccination titers (95% CI [0.97, 1.15], p<0.05), and LAIV-IIV with a 0.7 log_2_-unit increase (95% CI [0.47, 0.93], p<0.05), compared to IIV-IIV (**Figure S3C**).

### Viral antigen concentration impacted antibody titers across age groups

We next assessed 15,000 antibody titers from participants receiving distinct vaccine formulations: high-dose (Fluzone High-Dose), adjuvanted (Fluad), recombinant (Flublok), and standard-dose IIV as the reference. Using the original mixed-effects model, Fluzone High-Dose was associated with higher post-vaccination titers compared to standard-dose IIV (coefficient = 0.38 95% CI [0.30, 0.46], p<0.05) while neither Flublok nor Fluad showed significant improvements over the standard-dose formulation (**Figure 1**).

To examine this result by season, univariate analyses compared each formulation across all age groups and seasons, since these three factors had the largest effect sizes. In two seasons (2015, 2018), Fluzone High-Dose titers were significantly higher (absolute effect: ≥1.5x larger GMTs) than Fluzone in all age groups ≥60 years old. In six other seasons (2014, 2016, 2017, 2020, 2022, 2023), high dose was either comparable or only slightly better than standard dose in these age groups (<1.4x larger GMTs). In two seasons (2019, 2021), high dose elicited slightly but significantly smaller titers than standard dose in the 60-69 age range (0.65x smaller GMTs), with poorer performance also seen in the 70-79 age range in 2019 (**Figure 5**, **Figure S7**).

**Figure 5.**
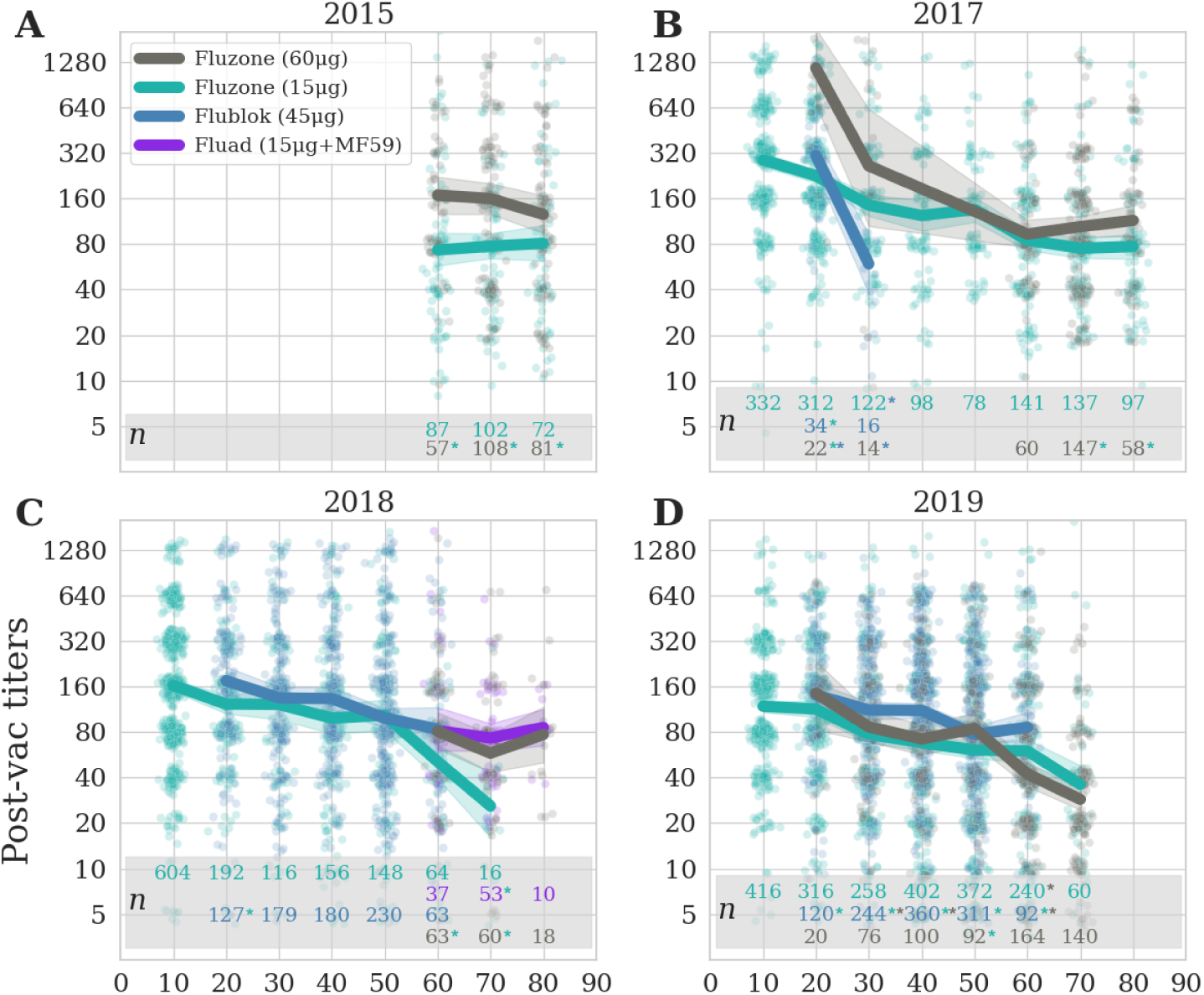
Antibody titers as a function of antigen dose. Post-vaccination titers for the (A) 2015, (B) 2017, (C) 2018, or (D) 2019 season. Lines show GMTs with 95% confidence intervals to standard dose Fluzone (15μg/antigen, teal), Fluad (15μg/antigen + adjuvant MF59, purple), Flublok (45μg/antigen, blue), or Fluzone High-Dose (60μg/antigen, grey), with participants dynamically binned by rounding their age down to the nearest decade so that each point represents at least 10 participants (**Methods**). The number of measurements (*n*) is color-matched for each age group and vaccine dose, with significantly higher titers denoted by an asterisk with the color of the poorer performing brand.

While Fluzone High-Dose is only approved for individuals ≥65 y.o., our studies included two clinical trials in 2017 and 2019 that assessed Fluzone High-Dose responses in younger age groups.^14,22^ In both seasons, standard- and high-dose vaccines often elicited comparable titers across all ages. A notable exception was that participants 20-29 y.o. in 2017 showed significantly higher titers with Fluzone High-Dose, suggesting that young adults may benefit from higher antigen content (**Figure 5B**).

Flublok also had mixed performance across seasons, eliciting smaller titers in 2016, 2017, and 2021 in at least one age group while eliciting slightly but consistently higher titers in 2018-2019 and dramatically higher titers in 2023 for 20-30 y.o. (**Figure S7**). In most seasons and age groups, Flublok titers were not significantly different from Fluzone. Fluad titers were only available in 2018 and 2023, yet in both seasons its titers were comparable to Fluzone High-Dose in all age groups.

### Multiple influenza vaccines elicit similar antibody titers for influenza A and B

We next expanded this analysis to compare the titers from multiple influenza vaccine brands to identify which formulations elicited the strongest antibody responses (**Figure 6**). Most vaccine brands elicited comparable titers, with ∼2x fold-change. However, a few notable exceptions were observed.

**Figure 6.**
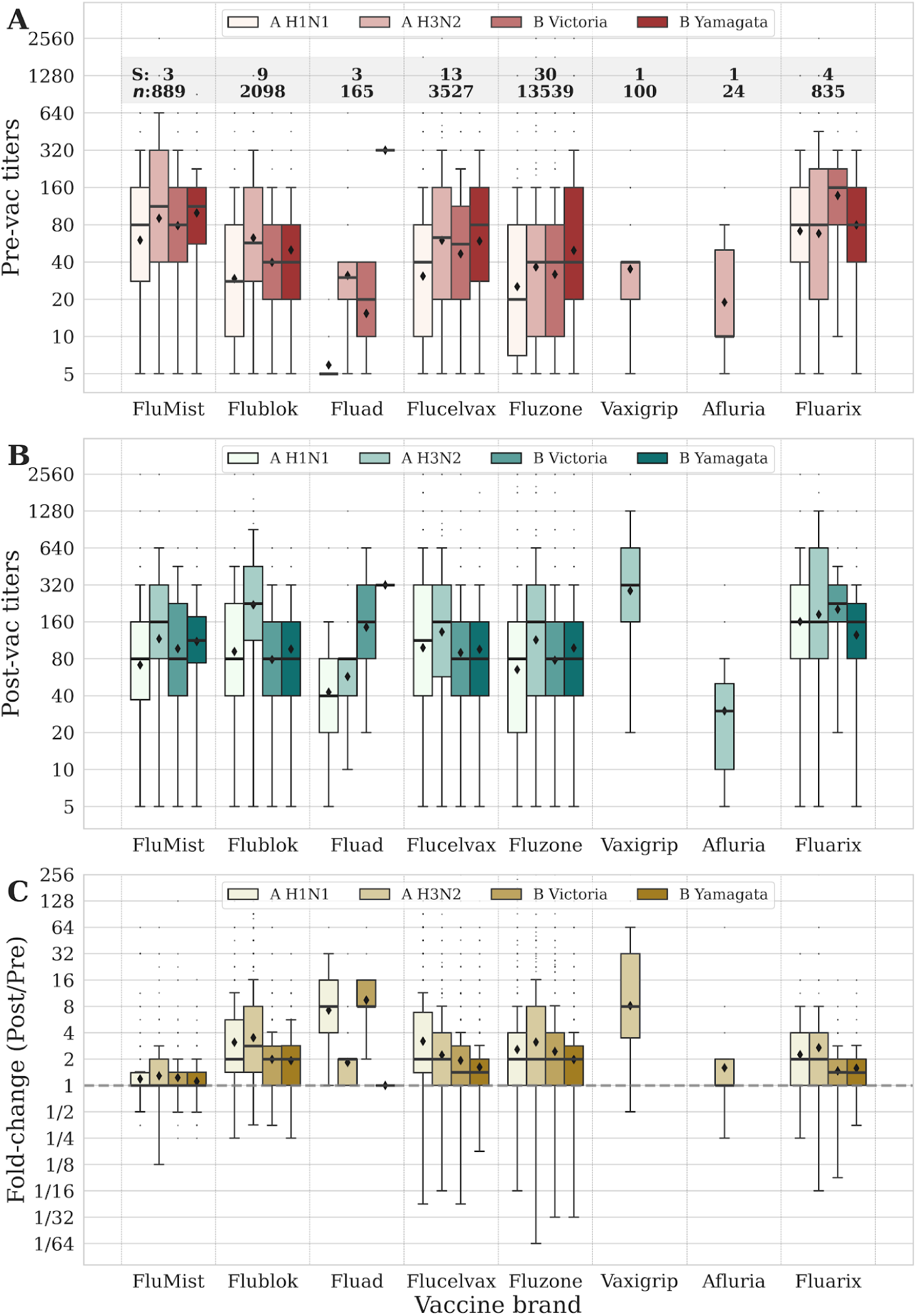
Unadjusted pre- and post-vaccination titers against each vaccine component and vaccine brand. Box plots comparing (A) pre-vaccination titer, (B) post-vaccination titer, and (C) fold-change across all seasons. The numbers in Panel A show the number of measurements (*n*) and number of studies (S) analyzed. The horizontal dashed line in Panel C indicates a fold-change of 1, corresponding to no change in titer. Box plots show the interquartile range, horizontal lines the medians, black diamonds the GMTs, and whiskers extend to 1.5 times the interquartile range.

First, FluMist showed no measurable increase in serum antibody titers with GMT fold-change consistently around 1 (**Figure 6C**), even though these participants had very low pre-vaccination titers, so any other vaccine would have likely elicited an excellent response. Second, Vaxigrip elicited an unusually high fold-change (∼8x GMT), although these responses were from a single study of adults in Vietnam that had never received a prior influenza vaccine. Third, Fluad showed elevated fold-changes for H1N1 and B Victoria, which appeared to be driven by unusually low pre-vaccination titers. The Fluad H3N2 pre-vaccination titers, which were more comparable to the other vaccine brands, resulted in the same ∼2x fold-change as other brands. Finally, the exceptionally high Fluad pre-vaccination titers against the B Yamagata lineage likely decreased its fold-change due to the antibody ceiling effect where larger baseline immunity leads to diminished fold-change (**Figure S8B**).^60,61^ When adjusted for pre-vaccination titers, Fluad was not statistically superior to standard-dose IIV (p = 0.41, **Figure 1**).

In summary, vaccine responses consistently showed 2-3x fold-change across all vaccine brands, with the key exceptions arising from: 1) FluMist that elicited little-to-no vaccine response, 2) large vaccine responses for adults receiving their first influenza vaccine, and 3) cases where pre-vaccination titers were substantially smaller/larger than normal, leading to larger/smaller fold-change.

### Minor sex differences were only seen against influenza A viruses in the elderly

Multivariate modeling found that males have log_2_ post-vaccination titers that are smaller by -0.1 units relative to females (**Figure 1**). This small effect size could not be seen in unadjusted univariate analyses, where there were minimal differences between males and females (**Figure 7A,B**). However, following prior work on frequently vaccinated individuals,^62^ a modest sex difference emerged in elderly individuals (age ≥65) who received multiple consecutive vaccinations. In this subgroup, females exhibited slightly higher antibody titers than males against influenza A viruses (**Figure 7C**). This difference only reached statistical significance during the third year of consecutive vaccination against H3N2 (p = 0.02, Mann-Whitney U test), yet in absolute magnitude women only had 1.5-fold larger titers. No significant difference was observed after 4 or 5 consecutive vaccinations, nor for H1N1 or either influenza B virus, suggesting that sex-based differences in antibody titers are very minimal, although age and repeated vaccination may reveal subtle, strain-specific effects.

**Figure 7.**
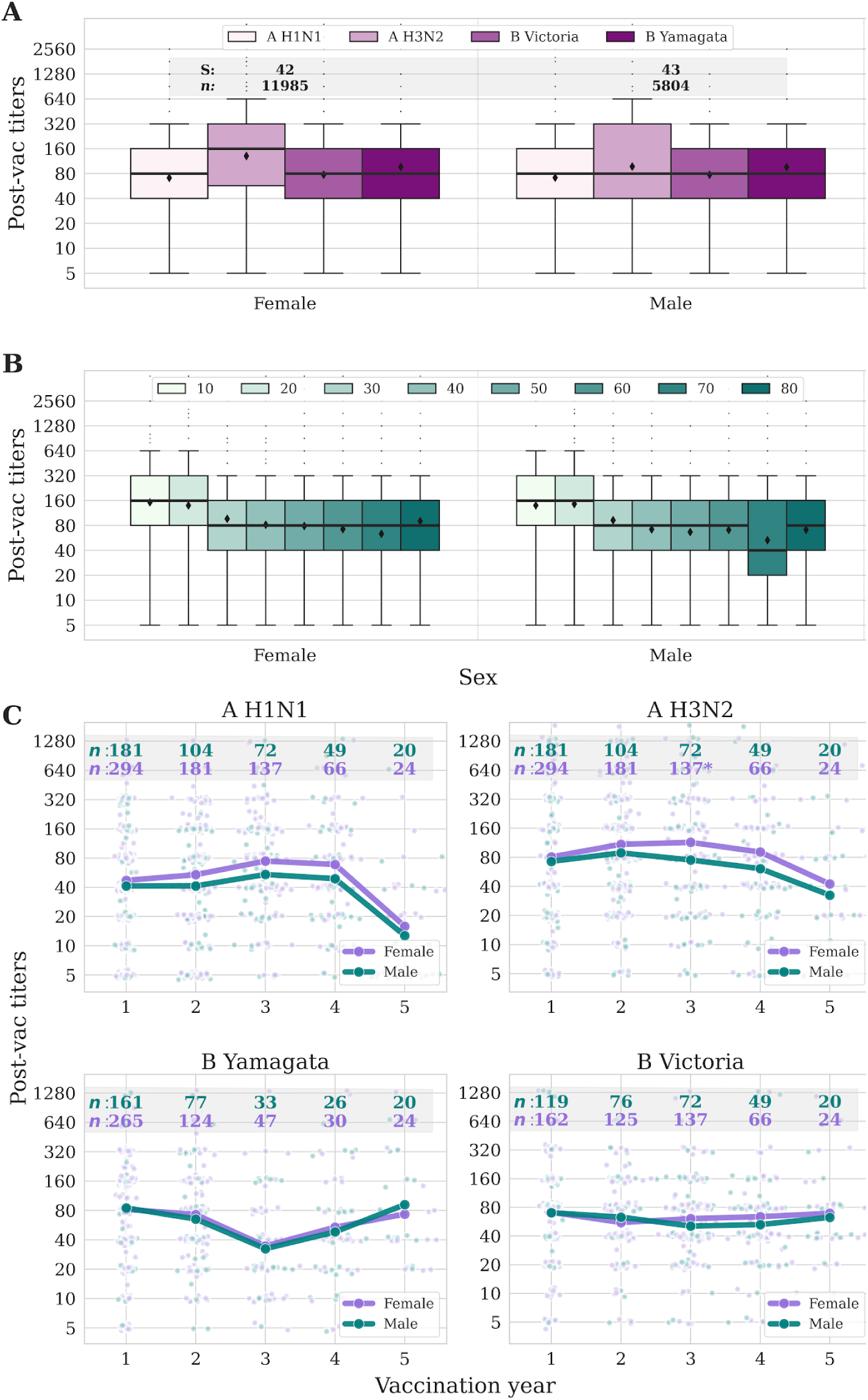
Unadjusted post vaccination antibody titers across sex. Post vaccination titers of males and females for each (A) vaccine component and (B) age. Box plots show the interquartile range, horizontal lines the medians, black diamonds the GMTs, and whiskers extend to 1.5 times the interquartile range. The numbers at the top show the amount of participants (*n*) and studies (S) included in this analysis. (C) Lines show GMT of post vaccination titers for groups of elderly (age ≥65) females and males that received 1 to 5 consecutive vaccinations. Colored numbers above each column denote the number of participants receiving their *k*^th^ vaccine, with stars indicating when the sex difference is significant (Mann-Whitney U rank test).

### Vaccinating later in the season elicited stronger antibody responses

To complement the determinants of immunity described above, we also used a neural network AI-guided discovery engine^63^ as a hypothesis generator to propose groups of features associated with the strongest and weakest post-vaccination titers or fold-change. While this engine identified four potential patterns, formal statistical analyses and verification were conducted using the univariate and multivariate methods described previously. The first two patterns involved age and pre-vaccination titers, which were described above and mildly affected antibody fold-change (**Figure S8A,B**). Interestingly, the combination of age and pre-vaccination titers led to larger effects that were not found through the mixed effects model, where participants <35 y.o. with low pre-vaccination titers achieved fold-change≈8x while older adults with low pre-vaccination titers only had 2-4x fold change (**Figure S9**).

The third AI-guided pattern focused on BMI, which in prior influenza and SARS-CoV-2 studies led to faster antibody waning and higher risks of hospitalization or death^64–67^ In our cohorts, BMI did not show a noticeable effect on the antibody response, suggesting that the AI method overfit on this parameter (**Figure S8C**).

The final pattern noted that the ∼10% of participants vaccinated late in the influenza season (between early January and mid-February) exhibited some of the highest post-vaccination antibody titers across the dataset. Extending this trend to encompass the full range of vaccine timing (September-February) showed a surprisingly consistent increase in fold-change, where geometric mean titers for individuals vaccinated in February were ∼2x larger than those vaccinated in September (**Figure 8A**).

**Figure 8.**
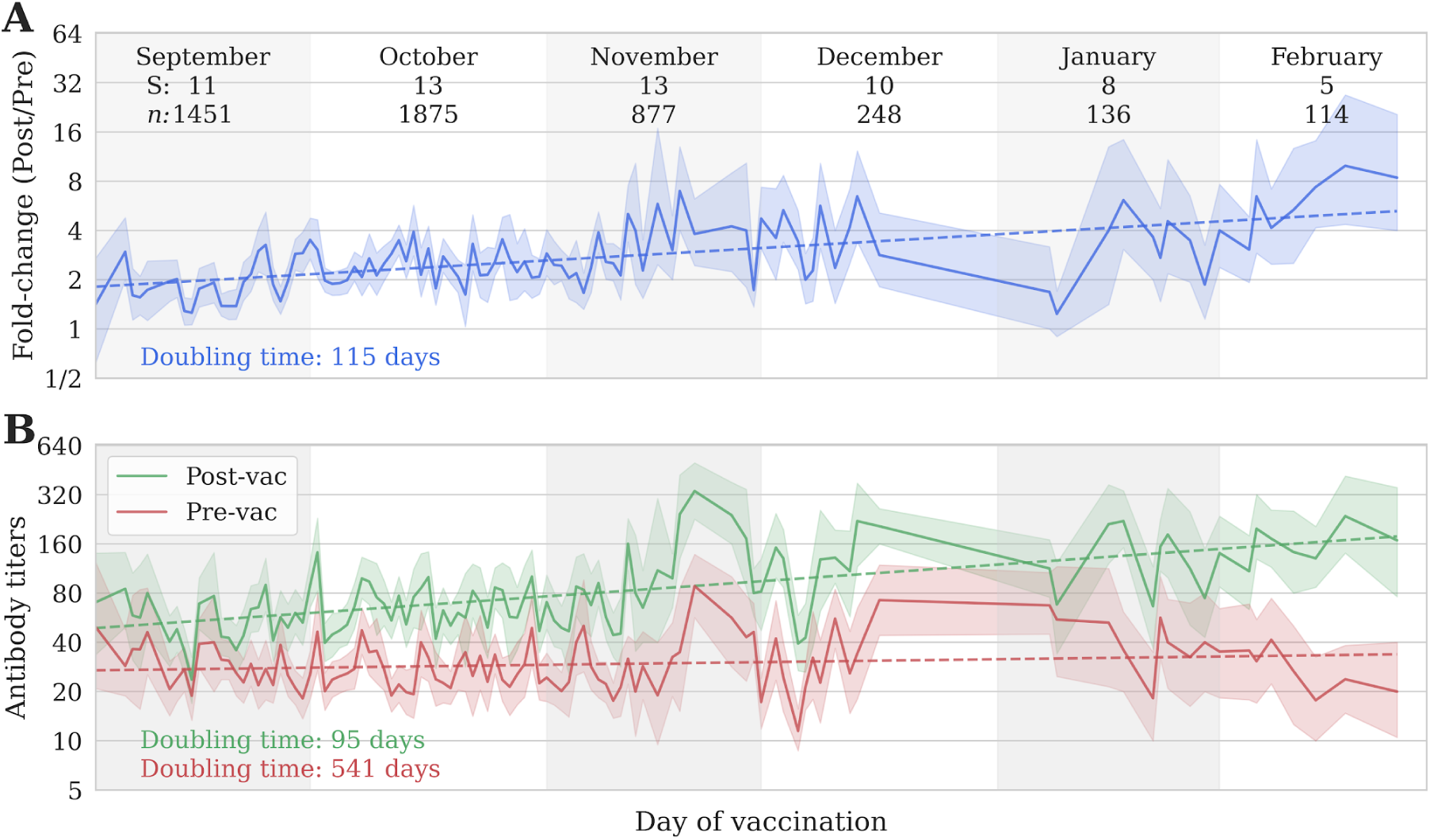
Unadjusted antibody titers through the season. (A) Fold-change and (B) pre-/post-vaccination titers of individuals vaccinated on different days of the year for northern hemisphere studies. The pre-vaccination, post-vaccination, and fold-change points are all shown at the *x* coordinate corresponding to the day of vaccination. Solid lines show the GMT of individuals at each day of the year with reported measurements. Shades represent the 95% confidence intervals. The number of measurements (*n*) and studies (S) are shown for each month.

Importantly, pre-vaccination titers remained largely stable, while post-vaccination titers progressively increased with later vaccination dates, suggesting that this effect is not caused by influenza infections priming the immune response (**Figure 8B**). These results held when stratified by age, season, or the time since last vaccination (**Figure S10A-C**). Increasing pre-vaccination titer led to a smooth and consistent decrease in fold-change (**Figure S10D**), as expected from the antibody ceiling effect that was clearly seen across studies (**Figure S11**). Thus, subsetting to participants with pre-vaccination titers≤10 revealed an even larger ∼5x increase in fold-change when vaccinated in February relative to September. Overall, these results suggest that vaccine timing can substantially affect immunogenicity.

To evaluate this effect through multivariate analysis, a *de novo* mixed effects model was run using data solely from the UGA studies (*n* = 9,100), which were the only datasets that recorded the date of vaccine administration. This analysis was therefore restricted to HAI titers run by a single lab, decreasing the likelihood of confounders from study design. After adjusting for all previously-described covariates in addition to vaccine timing, early-season vaccinations in October (p = 0.28) and November (p = 0.3) yielded post-vaccination responses statistically indistinguishable from September. However, later vaccinations administered in December (+0.29 log_2_ units), January (+0.47 log_2_ units), and February (+0.98 log_2_ units) generated significantly higher post-vaccination titers (p<0.05 for all), demonstrating that later vaccine administration increased post-vaccination antibody titers through both univariate and multivariate modeling (**Figure S3D**).

## Discussion

By integrating influenza serology data from more than 4,500 individuals across multiple seasons and study designs, this work provides an unusually broad view of how host, vaccine, and seasonal factors shape antibody responses. The following sections highlight the insights learned from analyzing influenza vaccine immunogenicity at scale.

### Seasonality and the impact from COVID-19

One of the largest effects on the influenza antibody response was season-to-season variation. It is generally expected that virus evolution and updates to the vaccine strains would yield highly variable antibody responses across seasons, since there were 14 updates to vaccine strains between 2014-2022 (**Table S2**). However, univariate and multivariate analyses showed that pre-vaccination titers were nearly flat from 2014 to 2019 and only dropped in the post-COVID-19 era (2021-2023). This decline in pre-vaccination immunity was most pronounced for H1N1, followed by more gradual decreases for H3N2 and B Victoria. This hierarchy of decline was unexpected, as it does not mirror known antigenic and genetic evolutionary rates: H3N2 undergoes the fastest antigenic drift and highest HA substitution rate,^68,69^ yet H1N1 titers declined most rapidly, reaching geometric mean titers of ≤15 as early as 2021. This suggests that background immunity is not solely determined by viral antigenic drift.

For post-vaccination titers, multivariate modeling showed that vaccination during 2023 elicited substantially lower titers than in the preceding decade. These poorer antibody responses in 2023 may have contributed to the large number and severity of infections in 2024, where the rate of hospitalizations was the highest observed since 2010.^70^

### Distinct response to B Yamagata

B Yamagata exhibited markedly different behavior than the other vaccine components, maintaining stable pre- and post-vaccination titer levels even after this lineage went extinct. This raises the question: Why did B Yamagata pre-vaccination titers not also decrease from 2021-2023 when there were few influenza cases?

Corroborating evidence of B Yamagata durability is seen from year two of the *Dynamics of the Immune Responses to Repeat Influenza Vaccination Exposures* study (DRIVE I), where the placebo group that received no vaccination in 2020 or 2021 nevertheless had stable HAI GMT≈40 for B Yamagata pre- and post-vaccination in both years (compared to GMT≲10 for H1N1 and H3N2 and GMT≈20 for B Victoria).^71^ Given that: 1) there were few influenza infections in Hong Kong during 2020-2021, 2) the inclusion criteria for this study required no vaccination during 2018 or 2019, and 3) vaccine coverage for adults in Hong Kong is generally low, these elevated B Yamagata titers have persisted for multiple years.

Since the datasets examined herein suggested that seasonal vaccines did do not elicit a durable antibody response at the population-level, we instead speculate that influenza infections elicit antibody titers that last multiple years, and that periodic reinfections helped maintain the pre-vaccination titers ≈40 observed before 2020. With the absence of influenza infections in 2020 and 2021 due to COVID-19 measures, this long-term immunity started to decline, revealing the baseline antibody levels. B Yamagata titers may have remained elevated because this lineage evolved more slowly both genetically^68^ and antigenically (**Figure S12**) than the other three vaccine components. Thus, each B Yamagata exposure closely matched the imprinted strain first encountered throughout life and elicited an especially strong and durable response (**Box 1**).

An intriguing point of comparison is the H2N2 subtype that circulated for 12 years (1957-1968), slowly evolving until it was replaced by H3N2. Individuals born during or before the 1950s still had inhibiting H2N2 A/Singapore/1/1957 antibodies (HAI GMTs≥40) more than 40 years after its extinction, while people born after 1968 had GTM = 5.^72–74^ These results further support that the first encounter to a slowly-evolving subtype leads to highly durable immunity, and this statement may hold for other pathogens beyond influenza.

Looking ahead, the elimination of B Yamagata provides a rare natural experiment to examine immune memory in the absence of both infection (since its extinction in 2021) and vaccination (since 2024 when vaccines became trivalent). Future studies should examine how long B Yamagata titers remain elevated in children versus adults, as well as from infection- or vaccination-primed individuals. Based on the H2N2 data, we predict that the B Yamagata titers will remain elevated for multiple decades.^73,74^

#### Box 1. Rationale for Decreasing Influenza Antibody Titers from 2020-2023 and B Yamagata Stability.

1. Pre-vaccination titers represent long-term immunity primarily conferred by infections.
  ○ In most vaccine studies, titers return to baseline at 1-year post-vaccination.
  ○ The two exceptions were adults given an influenza vaccine for the first time in their life,^71,75^ where fold-change was ≥8x at 1-month and ≥4x at 1-year post-vaccination in most subjects. These adults likely had prior influenza infections, suggesting that prior infections do not hinder this first durable vaccine response.
2. The lack of infections in 2020-2021 led to waning long-term immunity, revealing the baseline antibody titers against the current vaccine strains. Titers should therefore rise in the coming years.
3. B Yamagata titers have stayed high because this lineage mutates slower than H1N1, H3N2, and B Victoria.
  ○ We hypothesize that all B Yamagata variants are antigenically related, and that receiving any B Yamagata infection or vaccination yields an especially strong recall response.
  ○ In contrast, H1N1, H3N2, and B Victoria have all antigenically drifted, with the variants encountered in early life different from those in the 2020-2023 vaccines (that were used to assess pre-vaccination titers).

### Age effects

Multivariate modeling suggested that antibody titers in children steadily drop with age until 35 y.o., then hit a plateau between ages 35-64 y.o. before continuing to decline in elderly individuals 65-80 years old. While age effects have been extensively studied,^4,6–8,76–78^ an immunological transition in influenza immunity around age 35 has not been reported to our knowledge. In contrast, weaker antibody responses in the elderly have been extensively reported, yet the administration of Fluzone High-Dose successfully lifted the elderly antibody titers back to levels comparable with younger adults (**Figure S8A**).

### Most influenza vaccine brands behave similarly, except FluMist

Most vaccine brands elicited comparable antibody titers. One striking exception was FluMist, which aims to elicit mucosal responses (not examined in this work), but that consistently produced no measurable systemic antibody response across all cohorts (mean fold-change = 1.2x). Most adult studies similarly report that FluMist elicits no systemic antibody response.^20^ Indeed, FluMist has no known correlate of protection, and it was licensed based on vaccine efficacy and not immunogenicity.^20^ Studies in young children have been mixed, with some reporting antibody fold-changes ranging from 1-8x.^79–81^ Recent work in 2-5 y.o. receiving LAIV (with no prior influenza vaccinations) showed that only one third of children elicited a strong IgG response.^82^

### Skipping vaccination or the FluMist-skip

We also found a small and sporadic boost to antibody titers when participants did not vaccinate in the prior season. Between seven sets of consecutive seasons (2016-2017 to 2022-2023) and three age groups (children, adults, elderly), there were 8/15 instances where skipped vaccination (Skip-IIV) led to significantly higher post-vaccination titers than two consecutive vaccination (IIV-IIV), although the absolute increase in GMT was only 1.41x in these 8 seasons and 1.35x across all 15 cases. Year two data from the DRIVE I study corroborated these results, where groups receiving the Flublok (FB) vaccine or no vaccine (Skip) showed (post-vaccination GMT of FB-FB)/(post-vaccination GMT of Skip-FB) to H1N1 (0.73x), H3N2 (1.42x), B Vic (0.68x), or B Yam (0.84x) were all ≈1x.^71^ Future years of this study will investigate whether having more skipped years will yield a larger effect.

A distinct form of “skipped” vaccination involved administering FluMist followed by an inactivated influenza vaccine. Because FluMist does not typically elicit a systemic antibody response, this strategy could offer a similar immunological reset while still providing protection. To our knowledge, this vaccination sequence has not been formally tested, yet a subset of individuals in our cohorts followed it. Notably, in 2018-2019, adults who received FluMist followed by Fluzone exhibited significantly higher post-vaccination titers compared with those vaccinated with Fluzone alone (GMT: 166 vs 58). A similar pattern was observed in young individuals vaccinated with FluMist in 2019 and Flucelvax in 2020 (GMT: 340 vs 107), and further subsetting this group into younger (2-18 y.o., *n* = 44) and older (18-34 y.o., *n* = 40) age groups continued to show enhanced responses in each case with GMTs of 387 and 296, respectively. These results are also consistent with prior studies that also reported enhanced IIV responses following FluMist vaccination.^59,83^ It is important to note that only 24 individuals received the FluMist→Fluzone sequence and 84 received FluMist→Flucelvax, and that differences were not significant in young individuals in 2018-2019. Thus, these findings are suggestive but not conclusive.

### Sex differences

Across all cohorts, male sex led to a small but significant decrease in post-vaccination antibody titers. Some prior studies also showed modest sex differences where females had 1-2x larger titers than males,^62,84^ while other work found no difference in antibody titers but saw sex differences in ELISPOT or NAI responses.^85,86^ These subtle differences may reflect interactions between sex hormones, immune senescence, and repeated antigen exposure, but they do not appear to be a dominant determinant of influenza vaccine responsiveness.

### Time of Year Vaccinated

The impact of vaccine timing on the subsequent antibody response remains underreported and underexplored. Indeed, among our 64 datasets, the day of vaccination was only available in the 8 UGA studies. Yet across those vaccine responses, delayed vaccination in February led to ∼2x larger fold-change than early vaccination in September (or ∼5x larger fold-change for subjects with low baseline titers). Since pre-vaccination titers remained flat across all months, this effect was likely not caused by infections inflating baseline immunity. Previous studies noted that getting vaccinated too early in the season can increase the risk of infection due to within-season antibody waning,^87–89^ yet to our knowledge, increased titers from vaccinating later in the season have not been reported.

Mechanistically, we hypothesize that these changes in the immune response could be related to annual fluctuations in immune cells such as lymphocytes and neutrophils that prior reports have associated with daylength,^90^ or to increases in vaccine potency from storage. Although lymphocyte counts remain relatively stable from September through February, neutrophil counts show a marked increase, which may contribute to enhanced vaccine responses through their role in coordinating adaptive immune responses.^91^ Future work should investigate such mechanisms, and also explore this effect in southern hemisphere cohorts.

### Limitations

Analyses were performed on pooled data from multiple studies, providing the statistical power necessary to detect population-level immunological trends. However, this approach inherently introduces limitations regarding study design and assay heterogeneity.

First, while our multivariable mixed-effects model adjusted for cohort-level clustering and host traits, there may still be selection bias. For example, individuals that volunteered for a repeated-vaccination or long-term durability measurements may systematically differ in their underlying health status, healthcare engagement, or have more unrecorded prior vaccinations. Additionally, the majority of data analyzed were derived from cohorts in the United States, which may limit its generalizability to other populations.

Second, variations in laboratory protocols across the 64 pooled studies may have introduced systematic variability into the assay. While standard hemagglutination inhibition and microneutralization assays were utilized, specific operator interpretations and the use of complements like oseltamivir or ether were not documented in the original metadata. Additionally, comparing responses across egg-based and cell-based vaccines introduces potential bias regarding antigenic match to circulating viruses. Because the primary scope of this work was the strict evaluation of vaccine-induced antibody titers, extrapolating these results to absolute clinical protection against circulating, non-vaccine strains requires caution.

Third, administering a live attenuated influenza vaccine (LAIV) followed by an inactivated influenza vaccine (IIV) in the subsequent year is not a common clinical practice, inherently limiting the data available for this specific sequence. Compared to the robust sample sizes of the Skip-IIV group (*n* = 1763) and the IIV-IIV group (*n* = 5819), the LAIV-IIV group was relatively small (n = 164). When stratifying this cohort by season, the sample size in certain subgroups dropped to as low as *n* = 24.

Finally, analyzing immunological durability over multiple years relies on assuming follow-up measures represent fixed timepoints (*e.g.*, day 365 or day 730). Like all long-term longitudinal vaccine studies, this structure cannot fully account for exact sampling window variations or undocumented interim influenza exposures. An undocumented natural infection between day 30 and day 365 would artificially elevate apparent durability.

### Translational applications

The recent decline in pre-vaccination titers underscores the need to understand why antibody levels fall across the population. The remarkable durability of B Yamagata titers demonstrates that strong and lasting influenza immunity is possible, offering a benchmark for what effective influenza immunity could look like.

Vaccine timing emerged as a potential lever to partially improve post-vaccination titers, suggesting that prospective trials are needed to determine whether timing recommendations could be optimized to align with anticipated influenza circulation. The timing of influenza seasons can be inferred from prior seasons, near-real-time surveillance of the current season, or forecasts of hospitalizations in the coming weeks.^92–94^ Importantly, vaccinating early risks both smaller post-vaccination titers and within-season waning^89,95^, while delayed vaccination extends the window of unprotected exposure.

In addition, while skipped vaccinations slightly enhanced vaccine responses the following year in some age groups and some seasons, this effect was inconsistent and often mild, and it leaves the population unprotected during the skipped season. In contrast, 2 out of 3 cases where individuals were primed with FluMist in one season and given an inactivated vaccine (Fluzone or Flucelvax) the following season led to noticeable increases in the antibody response, suggesting that alternating across vaccine brands can yield better results. If some feature of FluMist durably enhances inactivated vaccines, that feature could potentially be incorporated into future vaccines.

Current and next-generation vaccine design often relies on introducing novel components (*e.g.*, adjuvants, neuraminidase) to test their efficacy. An alternative approach is to identify unifying principles of the most potent or durable immune responses observed in large cohorts, and then leverage these principles to improve the vaccine. For example, we found far more variability within than across age groups, with similar results for participants given different vaccine doses or with different frequencies of vaccination. Future efforts may benefit from extreme-phenotype analysis that intensively characterize individuals with the strongest and weakest responses to uncover additional actionable immunological mechanisms that govern vaccine immunogenicity.

## Methods

### Datasets Analyzed

Prior vaccine studies in **Table S1** have been previously described.^10,14,22,60,75,96–101^ The new vaccine studies introduced in this work were the 2019-2023 WHO-CBER studies run by the Center for Biologics Evaluation and Research within the FDA as part of the WHO influenza surveillance efforts using human sera from individuals vaccinated in the prior season.

For each of those WHO-CBER studies, the quadrivalent influenza vaccine from that season was administered (*e.g.*, 2019 WHO-CBER administered the 2019-20 influenza vaccine). H1N1 and influenza B were always measured by HAI, while H3N2 was measured using a neutralization assay. Metadata and vaccine dose are supplied in the supplementary information file, where repeatedly vaccinated individuals have the same *Base ID*. (Note that WHO-CBER participants were always unique across years, and hence they have no *Base ID* values). Post-vaccination samples were targeted for 28 days post-vaccination. As is standard procedure, infants under 13 months old were given two full vaccine doses 28 days apart, with their post-vaccination sample collected 28 days after the second dose.

For the WHO-CBER studies, vaccine strains were either cell-grown (for Flucelvax) or egg-grown (for all other vaccine brands). In the supplementary information file, cell-grown viruses have “_Cell”, “_MDCK”, or “_SIAT”, while all other variants were egg-grown. The vaccine formulation LAIV corresponds to the FluMist brand, while IIV corresponds to Fluzone, Vaxigrip, Afluria, and Fluarix.

### Vaccine Study Participants

The five new influenza vaccine studies presented in this work included 224 participants for 2019 WHO-CBER, 208 in 2020, 320 in 2021, 240 in 2022, and 560 in 2023. Each participant’s age, vaccine dose, and vaccine brand is provided in the supplementary information file, and no other metadata was collected. These studies used existing sera from prior vaccine studies, and hence they did not qualify as human subjects research and did not require an IRB.

### Analyzing Antibody Titers

Antibody titers from HAI or neutralization were collectively analyzed to quantify how much the antibody repertoire inhibits or neutralizes the influenza virus. Both assays are done using 2-fold dilutions, so antibody titers can equal 10, 20, 40… Titers below the lowest dilution (<10) were denoted by titer = 5. If HAI and neutralization were available for the same participant, only neutralization titers were kept.

In consecutive vaccination analysis (**Figure 3A**), each vaccine component is assessed separately, so that one participant can have up to four points in each column for every vaccine strain.

In the age analyses (**Figures 5, 7B**), points were first binned into decades (10-19, 20-29…80-89 years old), and then dynamically binned so that no bin had <10 points to ensure robust statistics. Any bin with fewer points was moved into its neighboring bin with the least number of points, with this process repeated until all bins had ≥10 points. Few participants were <10 or ≥90 years old, and these were discarded from the age analysis to ensure robust statistics, but these participants are included in the supplementary information file.

For the date-of-vaccination analyses (**Figure 8**), each of the 365 days of the year constituted one bin, and points were then dynamically binned until no bin had <10 points.

### Hemagglutination Inhibition and Neutralization Protocols

Hemagglutination inhibition assays were performed using standard methods. Each volume of serum was treated with 3 volumes of receptor destroying enzyme (Accurate Chemical, Westbury, NJ, USA) at 37°C overnight, which was then inactivated by heating in a 56°C water bath for 30 min. Six volumes of PBS were added to make the final serum concentration of 1:10. Sera were diluted in a series of 2-fold serial dilutions in 96-well plates (Thermo Fisher). An equal volume of influenza virus, adjusted to 8 hemagglutination units (HAU)/50µL diluted in 1x PBS, was added to each well of the plate. The plates were then mixed by gentle agitation, covered, and allowed to incubate for 30 minutes at room temperature. Freshly diluted (0.5%) turkey red blood cells (Lampire Biologicals) for H1N1 and B antigens, were added to all wells and the plates incubated for an additional hour. After incubation, the last dilution of sera that completely inhibited agglutination was recorded. The antibody titer was determined by taking the reciprocal dilution of the last well that contained non-agglutinated red blood cells.

An enzyme-linked immunosorbent assay–based microneutralization assay was performed to quantify antibody titer toward H3N2 viruses. The same sera from the hemagglutination inhibition assay, 1:10 diluted and treated with receptor destroying enzyme, were used for the neutralization assays. Sera were incubated with virus titrated to one hundred 50% tissue culture infectious dose at 37°C with 5% carbon dioxide for 1 hour, and then were added to 1.5×10^5^ MDCK-SIAT1 cells (Sigma). After overnight incubation, infected cells were detected using influenza A nucleoprotein-specific monoclonal antibodies (Millipore). Neutralization titers represent the reciprocal of the highest serum dilution resulting in ≥50% neutralization.

### Vaccine History

For the consecutive season analysis (**Figure 3**), each trajectory begins in the first season when a participant was enrolled in an influenza vaccine study and continues through subsequent seasons as long as the participant either participated in a vaccine study or self-reported as receiving an influenza vaccine. Self-reported vaccinations were recorded for up to three years prior to study enrollment. When a participant reported vaccination but did not participate in a vaccine study, antibody titers were unavailable and therefore excluded from the GMT calculations. In cases where an individual had two independent blocks of ≥3 consecutive seasons (*e.g.*, three seasons vaccinated, a skip year, and then three more seasons vaccinated), each block was treated as a separate trajectory defined by the first year of the series of consecutive vaccinations.

When linking responses across multiple influenza seasons (**Figure 4A,C**), antibody titers were always assessed against the same day 0 influenza strain, even if the vaccine strain was updated. For example, if a person is vaccinated in 2017 with H3N2 A/Hong Kong/4801/2014, and then in 2018 with H3N2 A/Singapore/INFIMH-16-0019/2016, we used their pre-vaccination antibody titer in 2018 against H3N2 A/Hong Kong/4801/2014 to quantify their day 365 response in 2017. In other words, durability relied on repeated participation in a vaccine study as well as that study measuring prior influenza variants when the vaccine strain changes.

### Multivariable linear mixed-effects model

To identify the determinants of vaccine-induced immunity while adjusting for their potential interdependence, we used the statsmodel library^102^ (version 0.14.5) to construct a multivariable linear mixed-effects model whose primary outcome was the log_2_-transformed post-vaccination titer. Fixed effects included the continuous log_2_ pre-vaccination titer, age (categorized into 5-year intervals with a 10-year-old baseline to capture non-linear effects), biological sex (reference: female), cumulative number of prior influenza vaccinations, standardized vaccine formulation (reference: standard-dose IIV), virus strain (reference: H1N1), study year (reference: 2010), assay type (reference: HAI), and antigen substrate (reference: cell-grown). The model was repeated with alternative bin widths of 1, 2, 3, and 10 years as sensitivity analysis to assess the effect of chronological age on post-vaccination antibody titers (**Figure S2**).

For example, the log_2_(post-vaccination titer) of someone aged 65-69 in 2023 measured against the H3N2 vaccine strain is expected to be -1.0 (effect of age) + -2.1 (effect of season) + 0.3 (effect of virus strain) relative to the references of both categories (subjects with age 10-14 in 2010 measured against the H1N1 vaccine strain). Additional features can be added linearly, while any unlisted features are assumed to match the references.

To account for unmeasured heterogeneity and clustering introduced by pooling diverse cohorts, the specific study dataset was included as a random intercept. We calculated the intra-class correlation from the model’s variance components to quantify the proportion of residual variance attributable to dataset-level batch effects versus individual-level variation.

To evaluate the impact of vaccination history, effect of skipping, and vaccine timing, we conducted targeted subanalyses. Because the mixed-effects models require complete data, we restricted these analyses to subjects with no missing data for any of the evaluated features.

### Statistical analysis

Statistical comparisons between independent measurement groups were performed using a conditional testing framework implemented in Python (version 3.13.9), utilizing the SciPy (version 1.16.3) and Pingouin (version 0.6.1) libraries. Prior to comparative analysis, the distribution of each group was evaluated for normality using the Shapiro-Wilk test, with a significance threshold of 0.05 utilized for all preliminary and primary statistical assessments. The selection of the primary analytical test followed a specific heuristic determined by both distributional characteristics and the total number of observations per group to ensure the most robust statistical inference.

For comparison sets where both groups satisfied the assumption of normality, or where the Central Limit Theorem was deemed applicable due to a large sample size (n≥100 per group), a Welch’s t-test was employed. This parametric approach was selected specifically for its robustness against unequal variances and the significantly unbalanced group sizes present in the study’s datasets. The magnitude of the difference for these parametric analyses was quantified using Hedges’ *g*, providing a bias-corrected estimate of effect size that accounts for the variation in sample sizes between groups. Conversely, in instances where the normality assumption was violated and sample sizes were insufficient for parametric approximation (n<100), the Mann-Whitney U test was utilized as a non-parametric alternative to assess distributional differences. For these non-parametric comparisons, the Rank-Biserial Correlation (r_rb_) was reported as the corresponding measure of effect size. To maintain statistical reliability, comparisons involving any group with fewer than three observations were excluded from the analysis. Finally, the p-values and 95% confidence intervals from our multivariable linear mixed-effects models were calculated by the statsmodels library using Wald Z-tests under the assumption of asymptotic normality, which is robust given the large sample size of our dataset (n>20,000).^102^

## Data and Code Availability

*For reviews:* All data is provided as a supplementary information file.

*When the manuscript is accepted*: All data and code will be made available at https://github.com/TalEinav/DeterminantsOfInfluenzaImmunity.

## Data Availability

All data produced in the present study are available upon reasonable request to the authors

## Acknowledgements

We especially thank the experimental groups who shared their data. Part of the data was provided by the Centers for Disease Control and Prevention/Agency for Toxic Substances and Drug Registry, who compiled data from the 2018-2019 CDC studies and the 2019-2020 Williams studies. The new vaccine studies presented herein were from the Food and Drug Administration’s human serology efforts as part of the annual WHO influenza strain selection. We hope this paper will inspire other groups to integrate their datasets for everyone’s benefit, and we always welcome pointers to new datasets. We further acknowledge Chris Brown, Brendan Flannery, Jason Hsiao, Hannah Stacey, and Katherine Williams for useful discussions.

This research was supported by the the National Institute of Allergy and Infectious Diseases (NIAID) of the National Institutes of Health (NIH) under the Computational Models of Influenza Immunity (U01 AI187062), LJI & Kyowa Kirin, Inc. (KKNA - Kyowa Kirin North America), and the Bodman family (TE).

This paper is an informal communication and represents the authors’ best judgment. The material in this paper does not bind or obligate FDA.

## Notes

### Competing Interest Statement

The authors have declared no competing interest.

### Author Declarations

Ethics committee/IRB of the Center for Biologics Evaluation and Research, U.S. Food and Drug Administration, waived ethical approval for this work, as the study used existing sera and previously collected data and did not constitute human subjects research.

### Summary of Updates

Incorporated mixed-effects models to account for vaccination timing and history. Added mixed-effects models to analyze pre-vaccination titers. Conducted a sensitivity analysis utilizing age binning. Expanded the discussion of study limitations. Clarified the role of the AI-guided discovery engine. Revised and refined the manuscript text to improve overall clarity and flow.

