## Supplementary information for "Modifiers of Influenza Vaccine Immunity from over Ten Years of Serological Data"

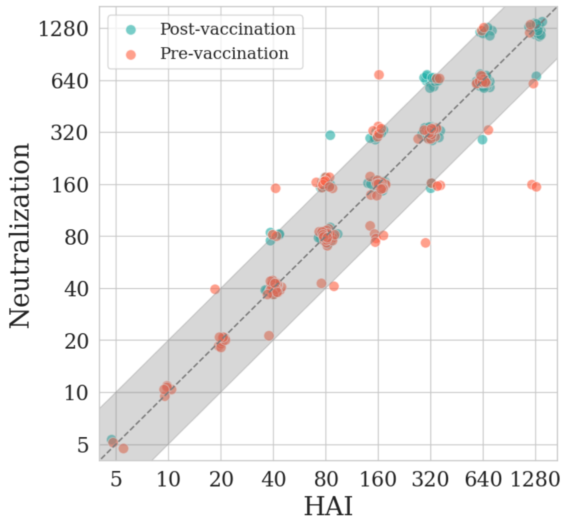


**Figure S1. Comparison between HAI and neutralization measurements.** Each point represents an individual with both HAI and neutralization titers measured against the same influenza variant either pre-vaccination (red) or post-vaccination (green). The dashed diagonal line indicates perfect agreement between the two assays. The shaded gray area denotes the ≈2-fold intrinsic error of each assay.

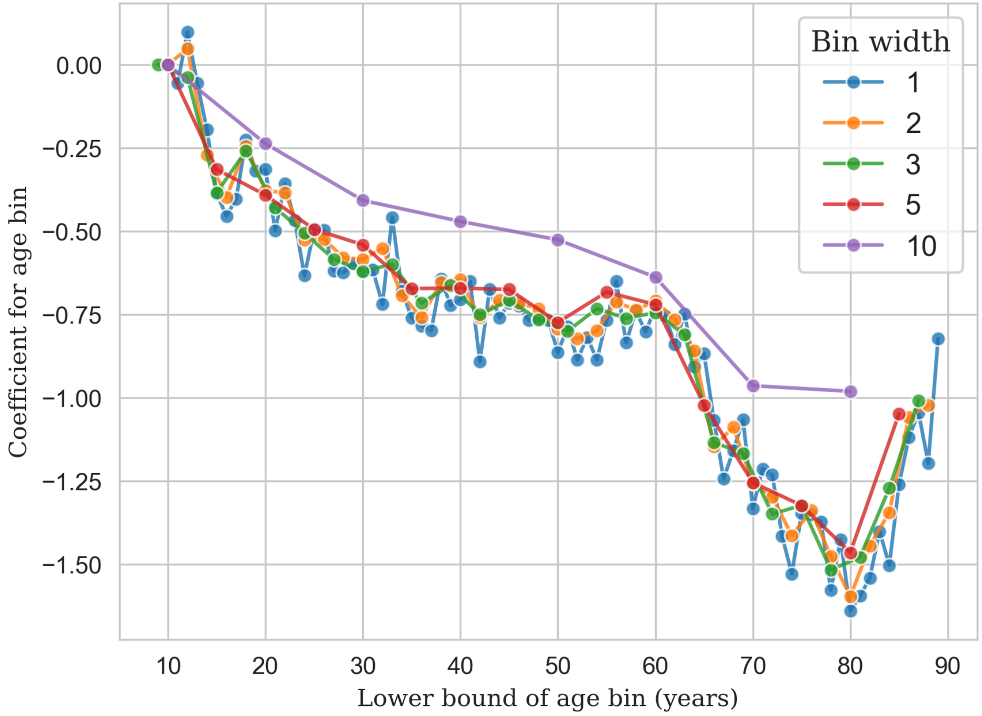


**Figure S2. Effect of age bin width on multivariable model coefficients.** The mixed-effects model from Figure 1 assessing post-vaccination titers was re-fit multiple times using age bins of 1, 2, 3, 5, or 10 years, with the resulting effect size shown for each model (*x* axis). All models were adjusted for pre-vaccination titer, number of previous vaccinations, vaccine brand, sex, season, passaging, assay, and virus.

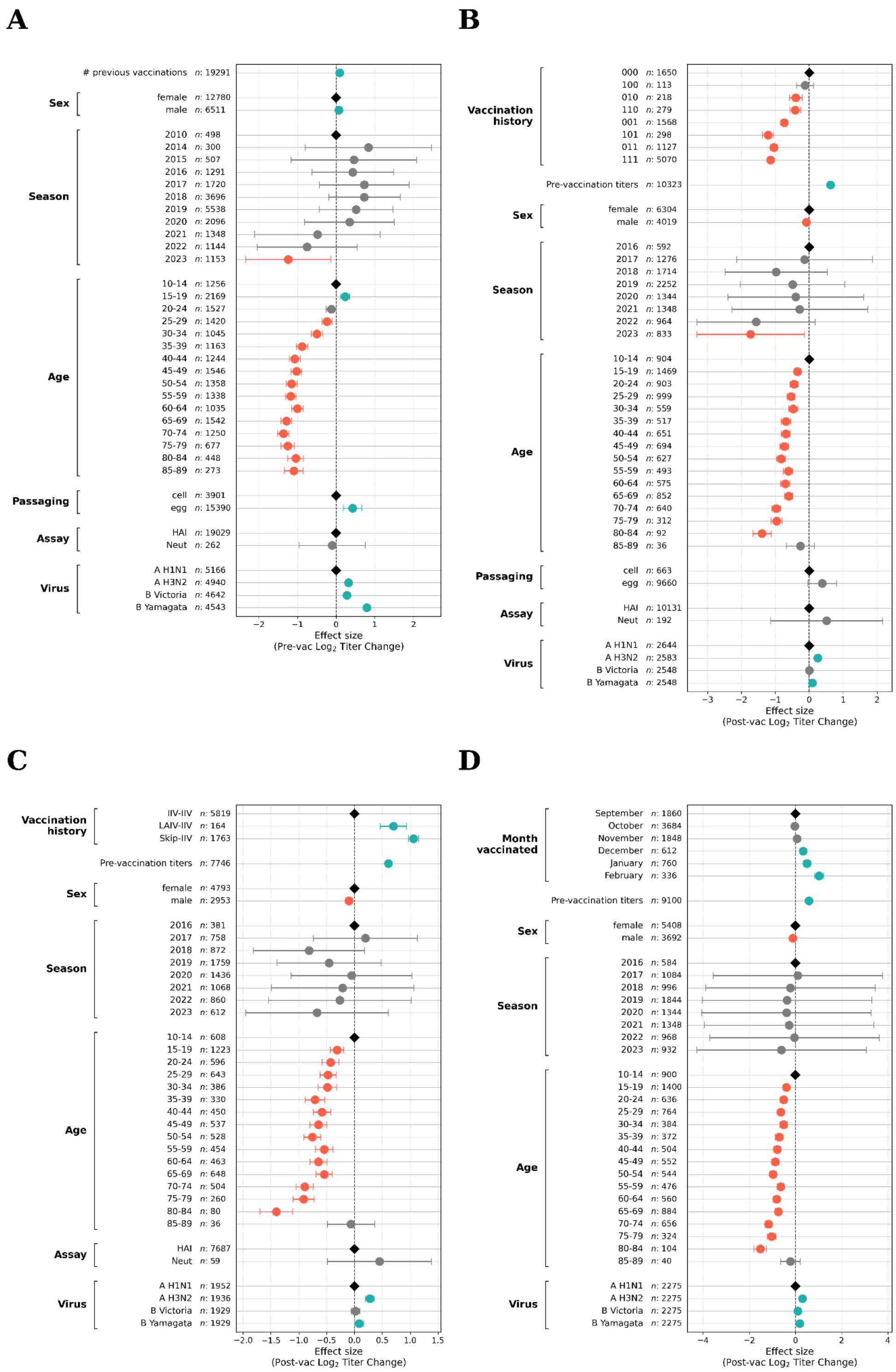


**Figure S3. Predictors of pre-vaccination titers, prior 3-season vaccination history, recent vaccination pattern, and month of vaccination through multivariate mixed-effects modeling.** Coefficients and 95% CI of log2(titer) effect size for each feature are shown on the *x* axis, with all covariates adjusted for shown on the *y* axis (**Methods**). (A) Predictors of pre-vaccination titer. (B) Effect of prior vaccination history on post-vaccination titer over the preceding three years (0 = not vaccinated, 1 = vaccinated, tuples represent vaccine history [3 years ago, 2 years ago, 1 year ago]). (C) Impact of skipping annual vaccination on post-vaccination titers. IIV-IIV, Skip-IIV, and LAIV-IIV denote an inactivated vaccine received in consecutive seasons, following a skipped season, or following FluMist the prior season, respectively. (D) Effect of month of vaccine administration on post-vaccination titer among UGA cohort participants. Teal, gray, and orange circles denote features that significantly increased, were non-significant, or significantly decreased titers, respectively, compared to the anchoring variable (black diamond, shown when relevant).

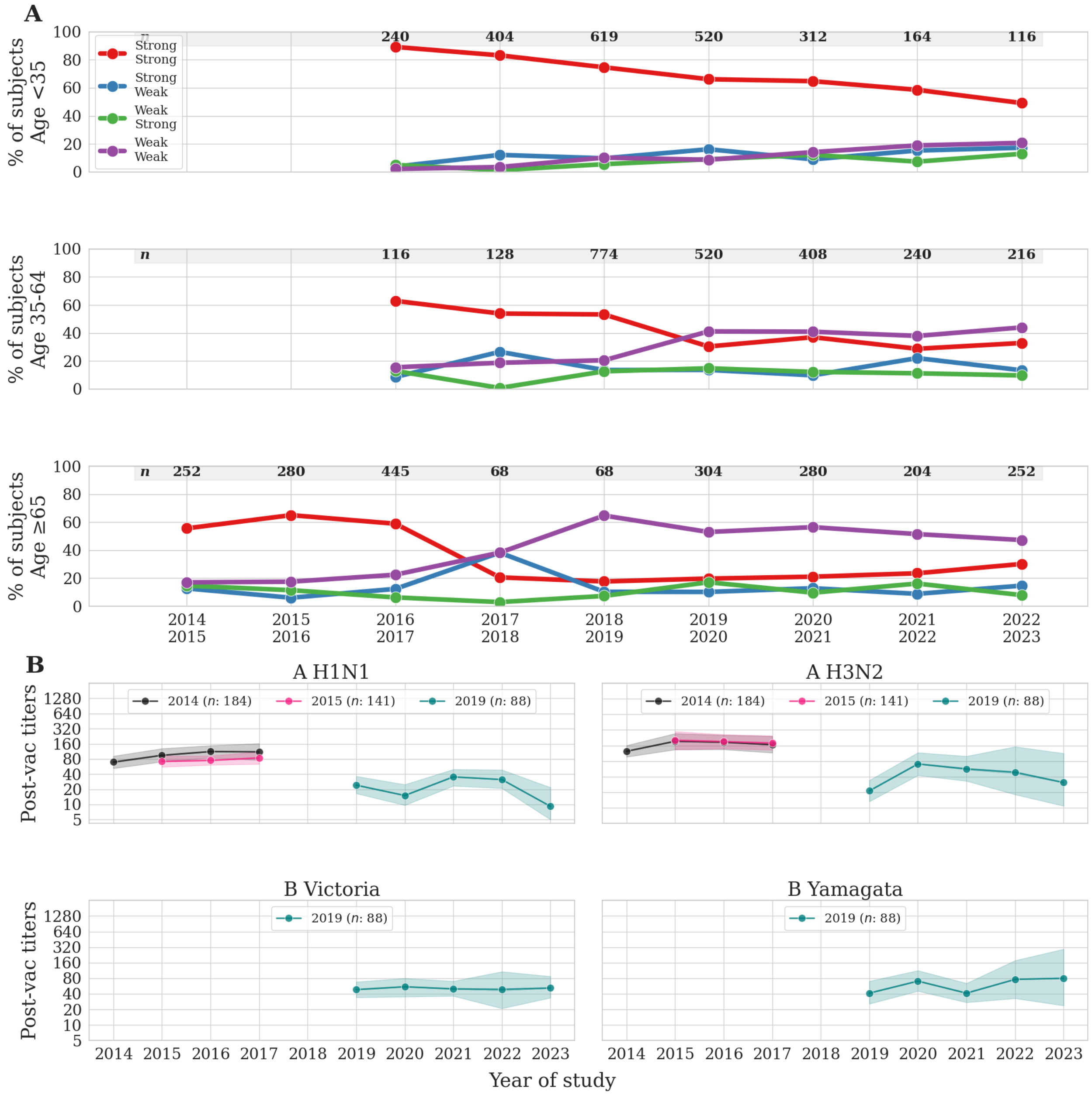


**Figure S4. Post-vaccination antibody titers in different immunological age groups for participants vaccinated in two consecutive seasons.** (A) Titers from participants clustered according to age were categorized as weak (≤40) or strong (>40) in two consecutive seasons. (B) GMT of participants aged ≥65 that were consecutively vaccinated in ≥3 consecutive seasons, subset by the first season of their consecutive vaccines (**Methods**). Lines show GMTs and 95% confidence intervals. The number of measurements (*n*) is shown in each panel.

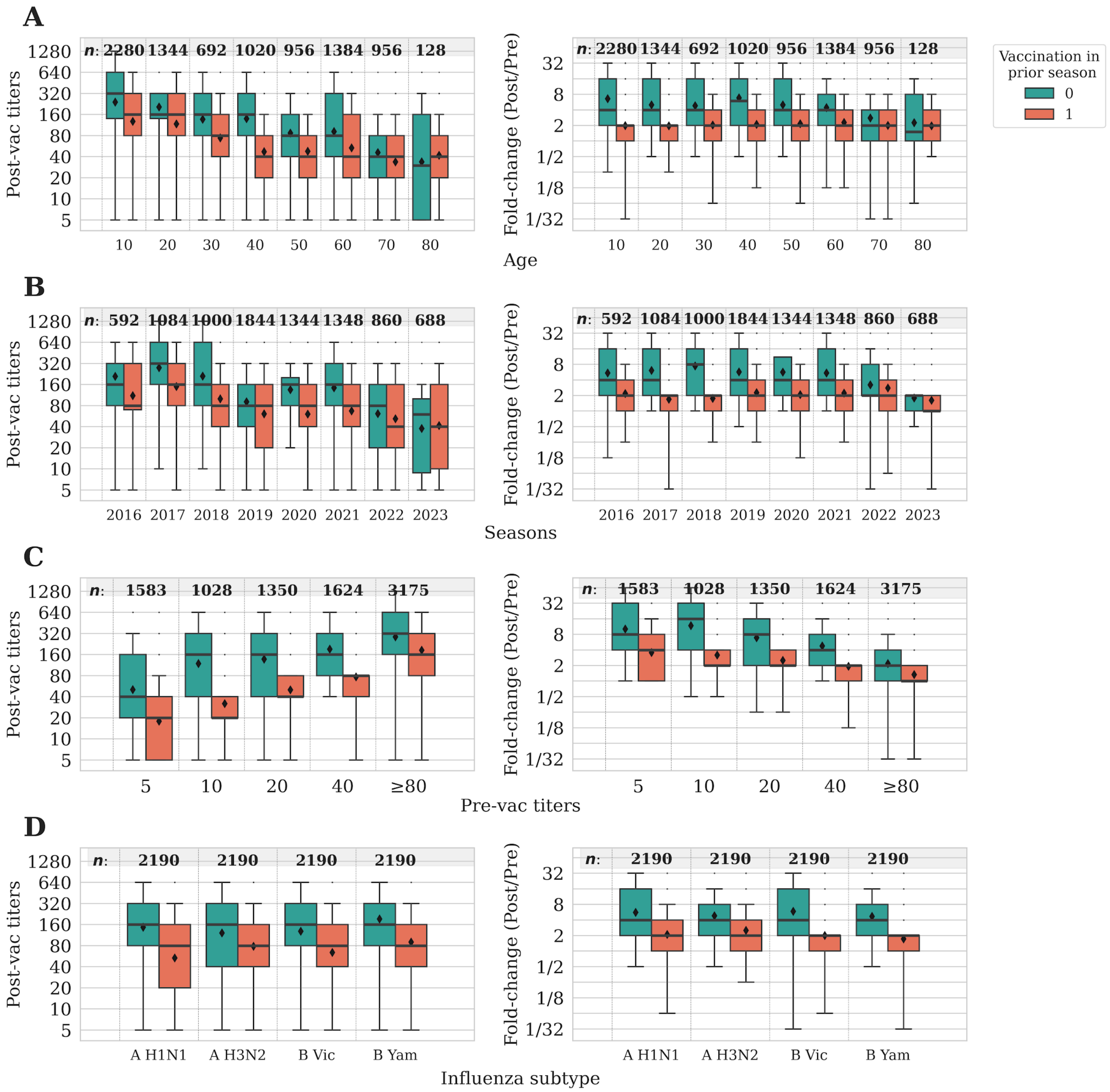


**Figure S5. Effects of vaccine history in post-vaccination titers and fold-change.** Stratification of vaccine titers of individuals that received (1) or did not receive (0) an influenza vaccine the previous season by (A) age, (B) season, (C) pre-vaccination titers, and (D) vaccine component. In each panel, the number of measurements is denoted by *n*.

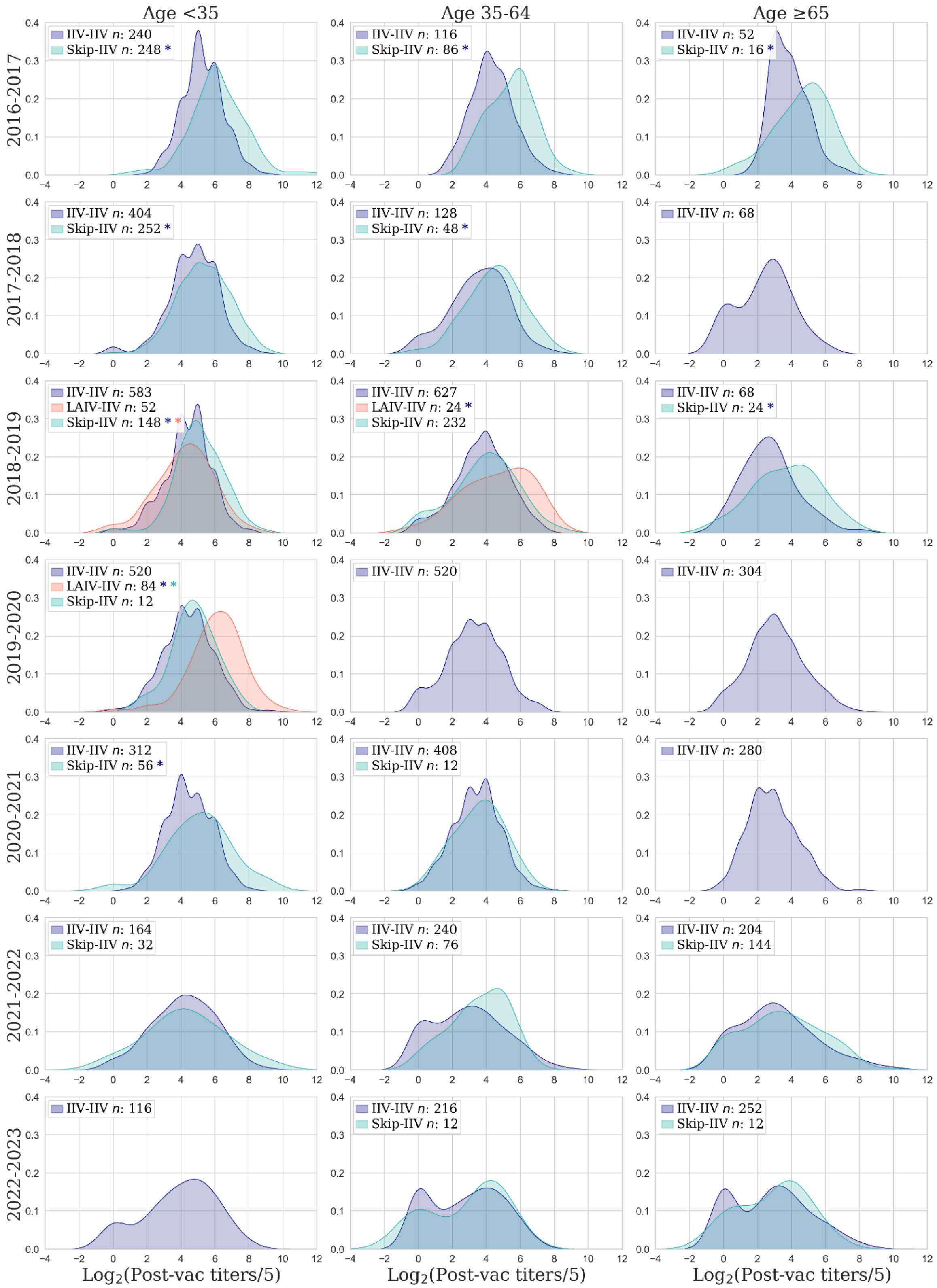


**Figure S6. Impact of skipping annual vaccination on antibody titers by seasons.** Density plots of post-vaccination titers for individuals aged<35, 35-64, and ≥65 years. Titers are shown for participants who received Fluzone or Flucelvax (IIV) in consecutive years (blue), Fluzone or Flucelvax following a skipped year (teal), or Fluzone or Flucelvax after using FluMist (LAIV) the previous season (red). A colored asterisk represents a statistically significant increase in post-vaccination titers over the group shown by that color using a Mann-Whitney U test (**Methods**).

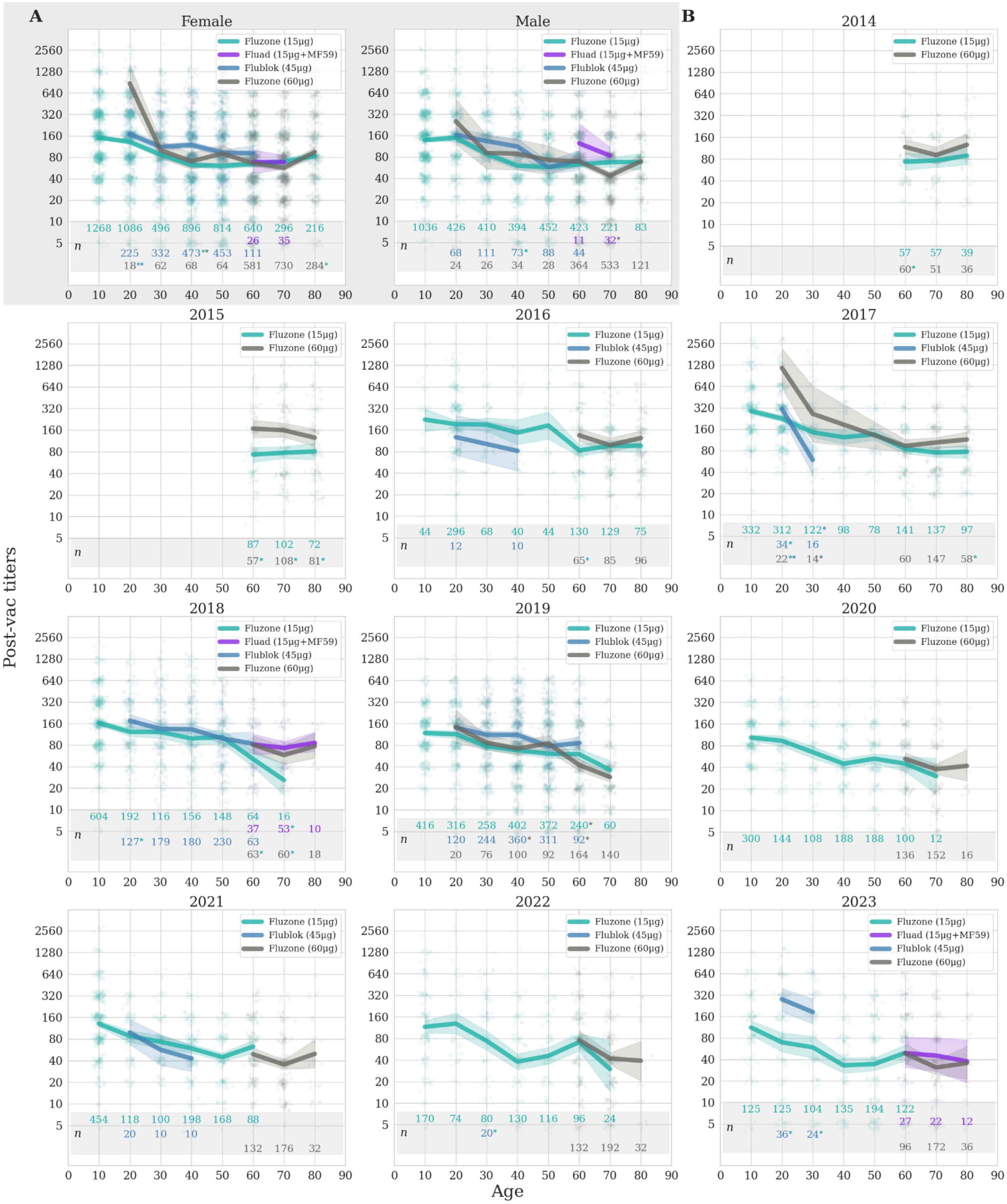


**Figure S7. Post-vaccination antibody titers for three antigen doses.** Antigen doses detailed by (A) sex, and (B) season. Lines show GMTs with 95% confidence intervals to Fluzone (15μg/antigen, teal),

Flublok (45µg/antigen, blue), Fluad (15µg/antigen + adjuvant MF59, purple), or Fluzone High-Dose (60µg/antigen, grey), with individuals dynamically binned by rounding their age down to the nearest decade so that each point represents at least 10 individuals (**Methods**). The colored numbers at the bottom show the number of individuals for each age group and vaccine brand. Significant improvements are marked by stars in the corresponding brand color.

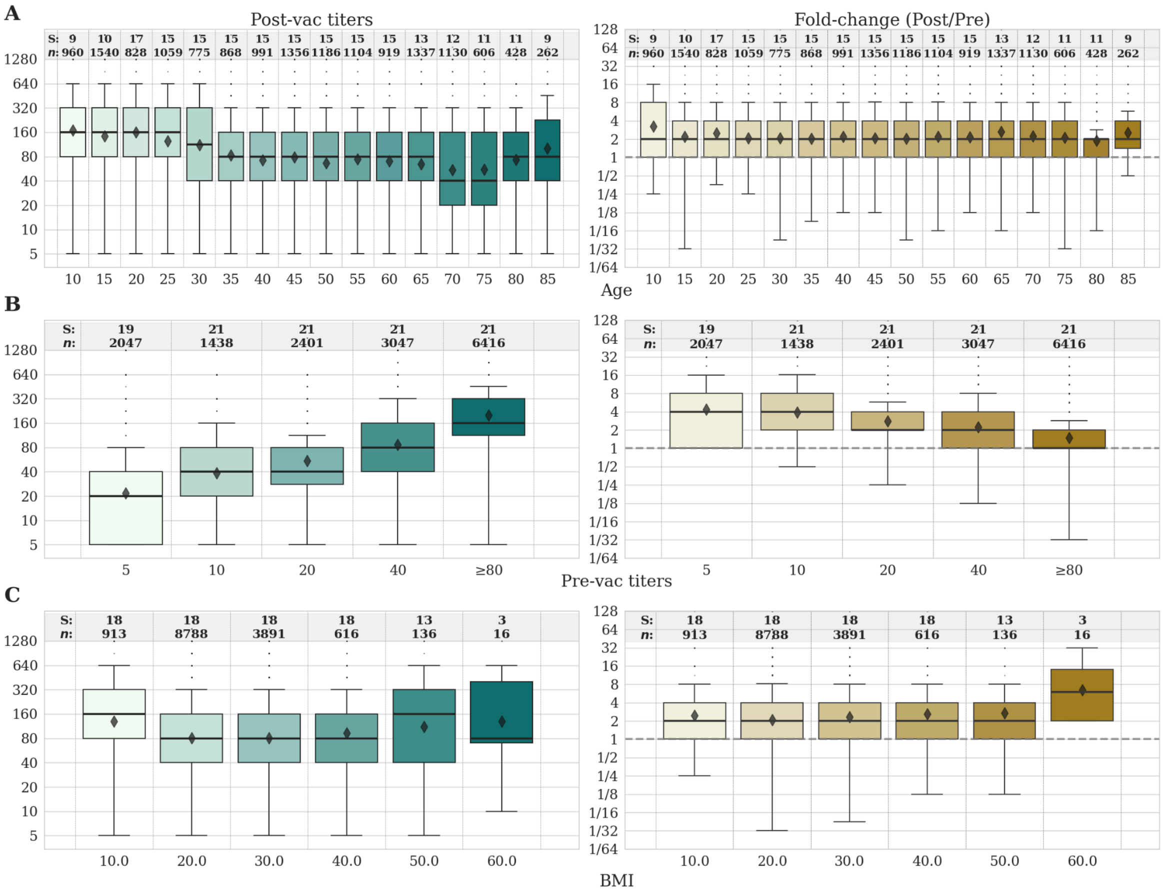


**Figure S8. Effects of host traits and baseline serology on antibody titers.** Post-vaccination titers and fold-change stratified by (A) age, (B) pre-vaccination titers, and (C) BMI. The numbers in the top panels show the number of measurements *(n)* and studies (S) analyzed. The horizontal dashed line in bottom panels indicates a fold-change of 1, corresponding to no change in titer. All box plots show the interquartile range, horizontal lines the medians, and black diamonds the GMTs. Whiskers extend to 1.5 times the interquartile range.

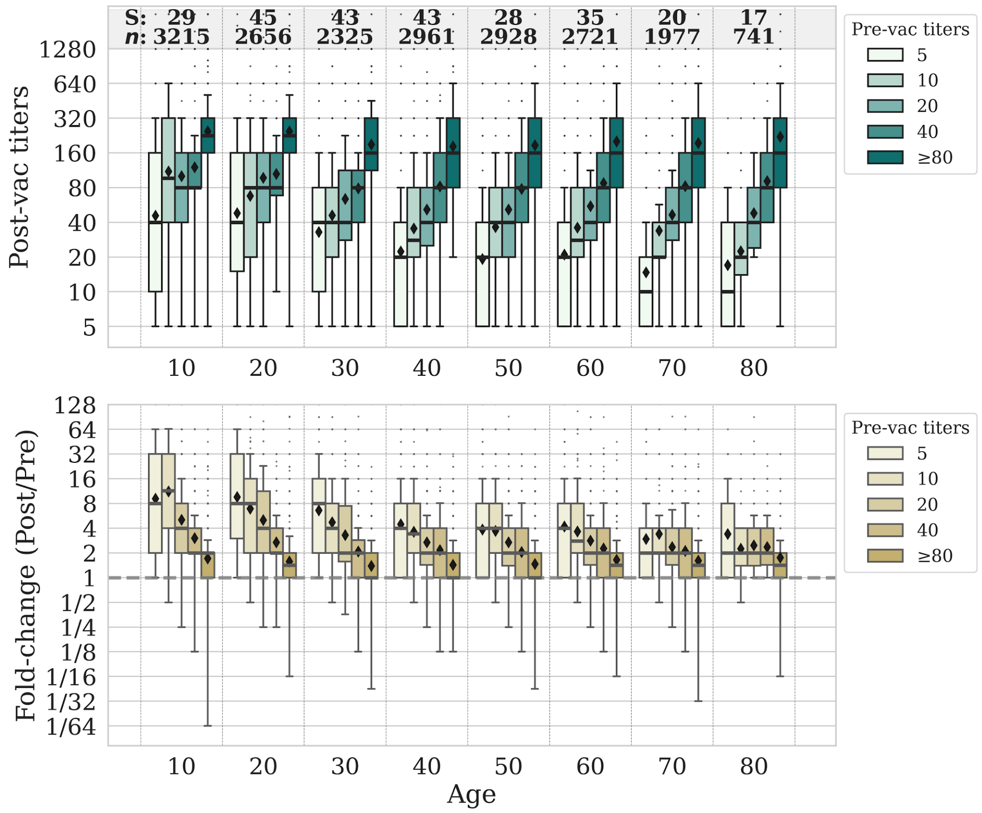


**Figure S9. Effects of age and pre-vaccination titers in post-vaccination titers.** Post-vaccination titers and fold-change stratified by age and pre-vaccination titers. The numbers in the top panel show the number of measurements (*n)* and studies (S) analyzed. The horizontal dashed line in the bottom panel indicates a fold-change of 1, corresponding to no change in titer. All box plots show the interquartile range, horizontal lines the medians, and black diamonds the GMTs. Whiskers extend to 1.5 times the interquartile range.

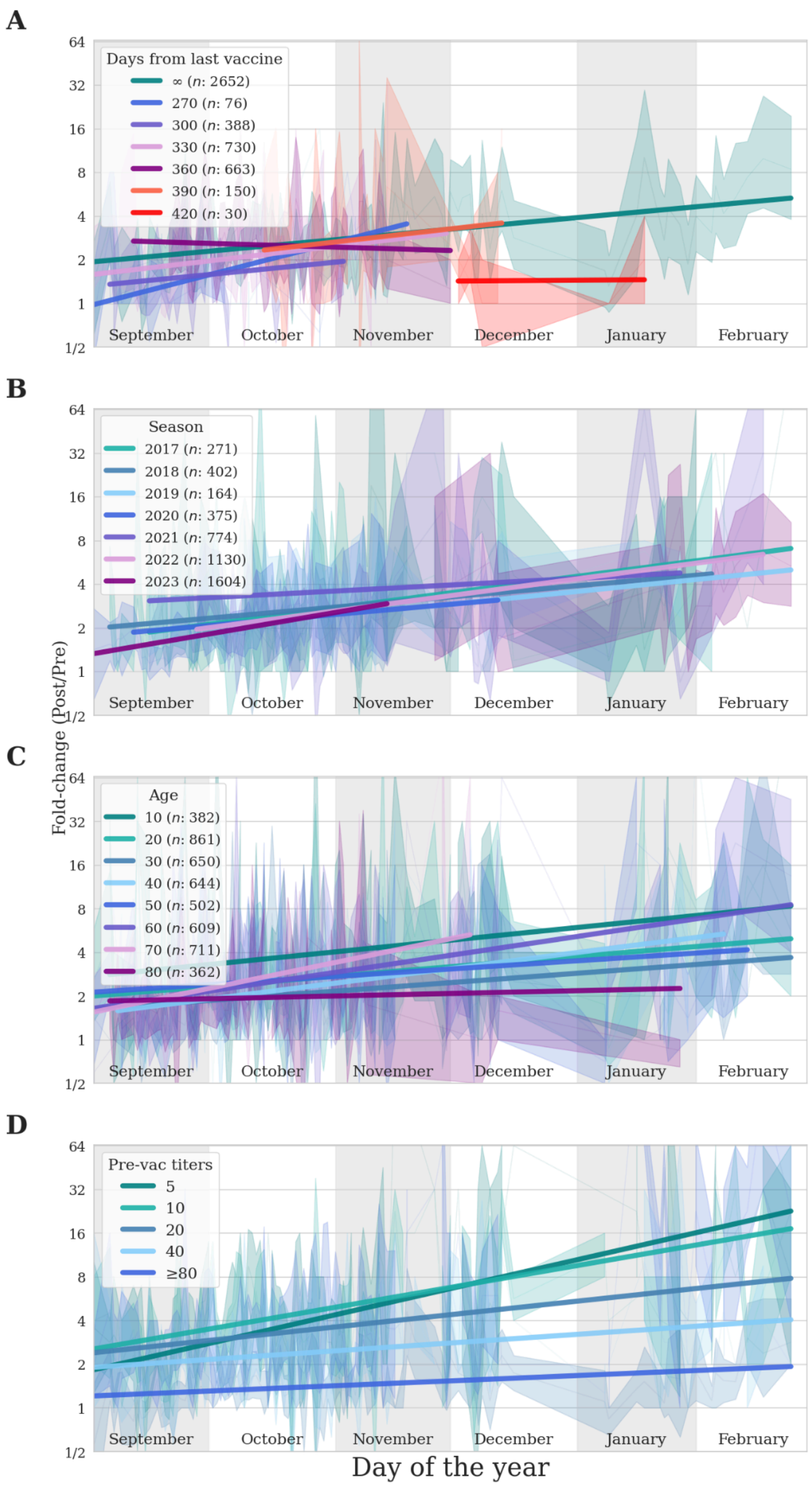


**Figure S10. Effect of vaccine timing on the antibody response.** Fold-change of individuals vaccinated on different days of the year stratified by (A) days since their last vaccination, (B) season vaccinated, (C) age, and (D) pre-vaccination titers. Solid lines show the GMT of individuals at each day of the year with reported measurements. Shades represent the 95% confidence intervals. Straight lines show the linear fit of each group.

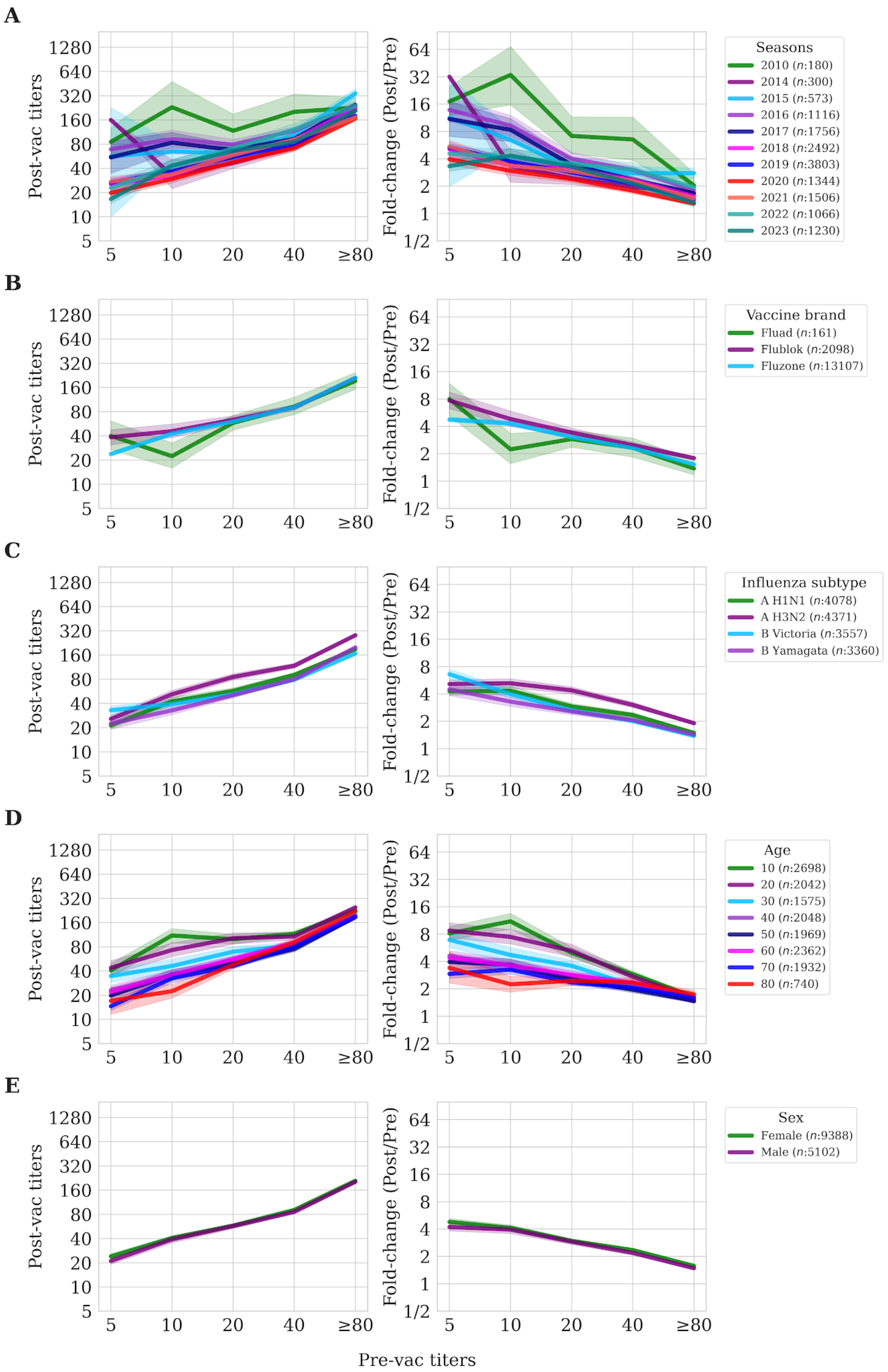


**Figure S11. Effects of pre-vaccination titers on post-vaccination titers and fold change.** pre-vaccination effects through (A) seasons, (B) vaccine brands, (C) vaccine component, (D) age and, (E) sex. Lines show GMTs with 95% confidence intervals with individuals dynamically binned by rounding their age down to the nearest decade so that each point represents at least 10 individuals (**Methods**).

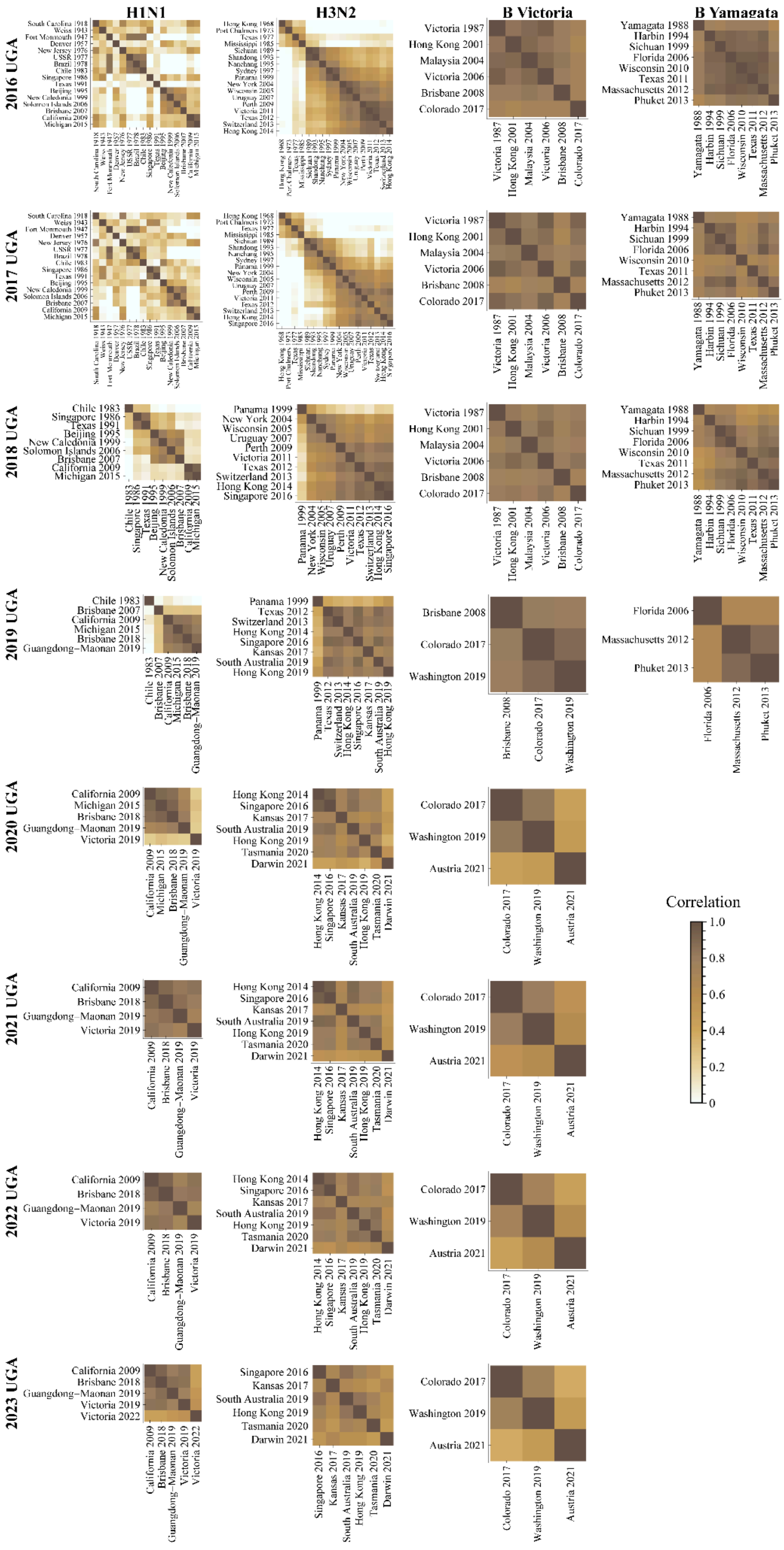
H1N1
H3N2
B Victoria
B Yamagata
2016 UGA
2017 UGA
2018 UGA
2019 UGA
2020 UGA
2021 UGA
2022 UGA
2023 UGA
Victoria 1987
Hong Kong 2001
Malaysia 2004
Victoria 2006
Brisbane 2008
Colorado 2017
Yamagata 1988
Harbin 1994
Sichuan 1999
Florida 2006
Wisconsin 2010
Texas 2011
Massachusetts 2012
Phuket 2013
Chile 1983
Singapore 1986
Texas 1991
Beijing 1995
New Caledonia 1999
Solomon Islands 2006
Brisbane 2007
California 2009
Michigan 2015
Panama 1999
New York 2004
Wisconsin 2005
Uruguay 2007
Perth 2009
Victoria 2011
Texas 2012
Switzerland 2013
Hong Kong 2014
Singapore 2016
Brisbane 2018
Guangdong−Maonan 2019
Kansas 2017
South Australia 2019
Hong Kong 2019
Washington 2019
Victoria 2019
Tasmania 2020
Darwin 2021
Austria 2021
Victoria 2022
Correlation
1.0
0.8
0.6
0.4
0.2
0

**Figure S12. Correlation between the combined pre- and post-vaccination titers of all variants measured in the 2016-2023 UGA studies.** Multiple variants were measured for nearly all vaccine components in each year, with the exception of 2020-2023 UGA that only measured the B Yamagata vaccine strain (not shown, since its correlation to itself would trivially be 1).

| Dataset | Year | Cohort | Vaccine Brand | HAI | Neutralization |
|---|---|---|---|---|---|
| CDC[103,104] | 2018 | | Fluarix | 75 | 25 |
| | 2018 | | Flublok | 147 | 46 |
| | 2018 | | Flucelvax | 216 | 72 |
| | 2018 | | Fluzone | 84 | 28 |
| | 2019 | | Flublok | 176 | 0 |
| | 2019 | | Flucelvax | 204 | 0 |
| | 2019 | | Fluzone | 480 | 0 |
| | 2018 | Repeaters | Fluarix | 276 | 92 |
| | 2018 | Repeaters | Flublok | 441 | 145 |
| | 2018 | Repeaters | Flucelvax | 621 | 205 |
| | 2018 | Repeaters | Fluzone | 246 | 82 |
| | 2019 | Repeaters | Flublok | 951 | 0 |
| | 2019 | Repeaters | Flucelvax | 915 | 0 |
| | 2019 | Repeaters | Fluzone | 244 | 0 |
| Crotty[105] | 2023 | | FluMist | 25 | 0 |
| | 2023 | | Afluria | 24 | 0 |
| Fox[75] | 2016 | HCW | Fluarix | 49 | 0 |
| | 2016 | Nam | Vaxigrip | 100 | 0 |
| Gouma[99] | 2017 | | Flublok | 0 | 50 |
| | 2017 | | Flucelvax | 0 | 52 |
| | 2017 | | Fluzone | 0 | 90 |
| Hinojosa[100] | 2015 | Hin_U | Fluzone | 31 | 0 |
| | 2015 | Hin_V | Fluzone | 41 | 0 |
| Kainth[10] | 2017 | | Fluzone | 88 | 0 |
| | 2018 | | Fluzone | 64 | 0 |
| | 2019 | | Fluzone | 20 | 0 |
| Kennedy[60] | 2010 | | Fluarix | 318 | 159 |
| | 2018 | | Fluad | 101 | 0 |

| | | | | | |
|---|---|---|---|---|---|
| | 2018 | | Fluzone | 110 | 0 |
| Loeb[98] | 2014 | | Fluzone | 312 | 0 |
| | 2015 | | Fluzone | 522 | 0 |
| | 2016 | | Fluzone | 510 | 0 |
| | 2017 | | Fluzone | 465 | 0 |
| Nakaya[96] | 2010 | ARM442 | Fluzone | 60 | 0 |
| | 2010 | ARM443 | Fluzone | 120 | 0 |
| UGA[14,101] | 2016 | | Fluzone | 616 | 0 |
| | 2017 | | Fluzone | 1144 | 0 |
| | 2018 | | Fluzone | 1056 | 0 |
| | 2019 | | Fluzone | 1992 | 0 |
| | 2020 | | Fluzone | 1488 | 0 |
| | 2021 | | Fluzone | 1520 | 0 |
| | 2022 | | Fluzone | 200 | 0 |
| | 2023 | | Fluzone | 176 | 0 |
| WHO-CBER (*This study*) | 2019 | | Fluzone | 92 | 92 |
| | 2019 | | Flucelvax | 20 | 20 |
| | 2020 | | Fluzone | 0 | 88 |
| | 2020 | | Flucelvax | 60 | 60 |
| | 2021 | | Fluzone | 80 | 80 |
| | 2021 | | Flucelvax | 60 | 60 |
| | 2021 | | Flublok | 20 | 20 |
| | 2022 | | Fluzone | 80 | 80 |
| | 2022 | | Flucelvax | 60 | 0 |
| | 2022 | | Flublok | 20 | 0 |
| | 2023 | | Fluzone | 200 | 100 |
| | 2023 | | Flucelvax | 120 | 20 |
| | 2023 | | Flublok | 40 | 20 |
| | 2023 | | Fluad | 40 | 20 |
| Williams[22] | 2019 | | FluMist | 392 | 0 |
| | 2019 | | Flucelvax | 400 | 0 |
| | 2020 | | FluMist | 472 | 0 |
| | 2020 | | Flucelvax | 448 | 0 |

| | | | | | |
|---|---|---|---|---|---|
| Zost[97] | 2016 | | Flublok | 22 | 0 |
| | 2016 | | Flucelvax | 26 | 0 |
| | 2016 | | Fluzone | 22 | 0 |

**Table S1. Every study included in the analysis.** The WHO-CBER studies are first presented in this work, while all other studies have been previously published.

| Vaccine Season | Recommended Vaccine Strains (Northern Hemisphere) | HA Amino Acid Changes from Prior Vaccine | % of US CDC Tested Viruses of this Subtype |
|---|---|---|---|
| 2023 | H1N1 A/Victoria/4897/2022 | 14 | 28.4% |
| | H3N2 A/Darwin/9/2021 | 0 | 66.9% |
| | B Victoria B/Austria/1359417/2021-like | 0 | 4.7% |
| | B Yamagata B/Phuket/3073/2013-like | 0 | 0% |
| 2022 | H1N1 A/Victoria/2570/2019 | 0 | 35.5% |
| | H3N2 A/Darwin/9/2021 | 10 | 62.1% |
| | B Victoria B/Austria/1359417/2021-like | 8 | 2.4% |
| | B Yamagata B/Phuket/3073/2013-like | 0 | 0% |
| 2021 | H1N1 A/Victoria/2570/2019 | 8 | 0.4% |
| | H3N2 A/Cambodia/e0826360/2020 | 16 | 99.0% |
| | B Victoria B/Washington/2/2019-like | 0 | 0.6% |
| | B Yamagata B/Phuket/3073/2013-like | 0 | 0% |
| 2020 | H1N1 A/Guangdong-Maonan/SWL1536/2019 | 9 | 27.8% |
| | H3N2 A/Hong Kong/2671/2019 | 18 | 38.9% |
| | B Victoria B/Washington/2/2019-like | 7 | 18.5% |
| | B Yamagata B/Phuket/3073/2013-like | 0 | 14.8% |
| 2019 | H1N1 A/Brisbane/2/2018 | 8 | 36.0% |
| | H3N2 A/Kansas/14/2017 | 16 | 21.3% |
| | B Victoria B/Colorado/6/2017-like | 0 | 38.9% |
| | B Yamagata B/Phuket/3073/2013-like | 0 | 3.9% |
| 2018 | H1N1 A/Michigan/45/2015 | 0 | 54.3% |
| | H3N2 A/Singapore/INFIMH-16-0019/2016 | 6 | 41.7% |
| | B Victoria B/Colorado/6/2017-like | 7 | 2.5% |
| | B Yamagata B/Phuket/3073/2013-like | 0 | 1.5% |
| 2017 | H1N1 A/Michigan/45/2015 | 18 | 10.2% |
| | H3N2 A/Hong Kong/4801/2014 | 0 | 57.4% |

| | | | |
|---|---|---|---|
| | B Victoria B/Brisbane/60/2008-like | 0 | 3.6% |
| | B Yamagata B/Phuket/3073/2013-like | 0 | 28.8% |
| 2016 | H1N1 A/California/7/2009 | 0 | 2.2% |
| | H3N2 A/Hong Kong/4801/2014 | 10 | 75.7% |
| | B Victoria B/Brisbane/60/2008-like | 0 | 6.4% |
| | B Yamagata B/Phuket/3073/2013-like | 0 | 15.7% |
| 2015 | H1N1 A/California/7/2009 | 0 | 57.1% |
| | H3N2 A/Switzerland/9715293/2013 | 8 | 13.7% |
| | B Victoria B/Brisbane/60/2008-like | 0 | 9.2% |
| | B Yamagata B/Phuket/3073/2013-like | 11 | 20.0% |
| 2014 | H1N1 A/California/7/2009 | 0 | 0.2% |
| | H3N2 A/Texas/50/2012 (A/Victoria/361/2011-like) | 0 | 83.2% |
| | B Victoria B/Brisbane/60/2008-like | 0 | 4.6% |
| | B Yamagata B/Massachusetts/2/2012-like | 0 | 11.9% |
| 2010 | H1N1 A/California/7/2009 | 117 | 28.1% |
| | H3N2 A/Perth/16/2009 | 10 | 45.8% |
| | B Victoria B/Brisbane/60/2008-like | 0 | 24.4% |
| | B Yamagata (Not in vaccine) | *N/A* | 1.5% |

**Table S2. Vaccine information for the seasons analyzed in this work.** 1st-2nd columns: North hemisphere vaccine strains. Throughout this text, “2010” refers to the 2010-11 season, “2014” the 2014-15 season, and so on. 3rd column: Number of HA mutations between each vaccine strain and the prior season’s vaccine strain. 4th column: Percent of circulating viruses from each season that antigenically matched to the vaccine strain (≤8x difference between the HAI of the circulating strain and the homologous virus using serum from ferrets infected with prior vaccine strains, taken from CDC’s Weekly Influenza Activity Reports).
